# Genome-wide association and Mendelian randomisation analysis among 30,699 Chinese pregnant women identifies novel genetic and molecular risk factors for gestational diabetes and glycaemic traits

**DOI:** 10.1101/2023.11.23.23298974

**Authors:** Jianxin Zhen, Yuqin Gu, Piao Wang, Weihong Wang, Shengzhe Bian, Shujia Huang, Hui Liang, Mingxi Huang, Yan Yu, Qing Chen, Guozhi Jiang, Xiu Qiu, Likuan Xiong, Siyang Liu

**Affiliations:** Central Laboratory, Shenzhen Baoan Women’s and Children’s Hospital, Shenzhen, Guangdong, China; School of Public Health (Shenzhen), Shenzhen Campus of Sun Yat-sen University, Shenzhen, Guangdong, China; Division of Birth Cohort Study, Guangzhou Women and Children’s Medical Center, Guangzhou Medical University, Guangzhou, China; Shenzhen Key Laboratory of Birth Defects Research, Shenzhen, Guangdong, China; Department of Obstetrics, Shenzhen Baoan Women’s and Children’s Hospital, Shenzhen, Guangdong, China; Department of Pharmacy, Shenzhen Baoan Women’s and Children’s Hospital, Shenzhen, Guangdong, China; Department of Women’s Health, Provincial Key Clinical Specialty of Woman and Child Health, Guangzhou Women and Children’s Medical Center, Guangzhou Medical University, Guangzhou, China

**Author notes:** Corresponding authors: Siyang Liu, Likuan Xiong. Jianxin Zhen and Yuqin Gu contributed equally to this study.

**Keywords:** Genetic risk factors, Molecular risk factors, Genome-wide association study, Gestational diabetes, Mendelian randomisation

## Abstract

**Aims/hypothesis:** Gestational diabetes mellitus (GDM) is the most common disorder in pregnancy; however, its underlying causes remain obscure. This study aimed to investigate the genetic and molecular risk factors contributing to GDM and glycaemic traits.

**Methods:** We collected non-invasive prenatal test (NIPT) sequencing data along with four glycaemic and 55 biochemical measurements from 30,699 pregnant women during a 2 year period at Shenzhen Baoan Women’s and Children’s Hospital in China. Genome-wide association studies (GWAS) were conducted between genotypes derived from NIPTs and GDM diagnosis, baseline glycaemic levels and glycaemic levels after glucose challenges. In total, 3317 women were diagnosed with GDM, while 19,565 served as control participants. The results were replicated using two independent cohorts. Additionally, we performed one-sample Mendelian randomisation to explore potential causal associations between the 55 biochemical measurements and risk of GDM and glycaemic levels.

**Results:** We identified four genetic loci significantly associated with GDM susceptibility. Among these, *MTNR1B* exhibited the highest significance (rs10830963-G, OR [95% CI] 1.57 [1.45, 1.70], *p*=4.42×10^-29^), although its effect on type 2 diabetes was modest. Furthermore, we found 31 genetic loci, including 14 novel loci, that were significantly associated with the four glycaemic traits. The replication rates of these associations with GDM, fasting plasma glucose levels and 0 h, 1 h and 2h OGTT glucose levels were four out of four, six out of nine, 10 out of 11 five out of seven and four out of four, respectively. Mendelian randomisation analysis suggested that a genetically regulated higher lymphocytes percentage and lower white blood cell count, neutrophil percentage and absolute neutrophil count were associated with elevated glucose levels and an increased risk of GDM.

**Conclusions/interpretation:** Our findings provide new insights into the genetic basis of GDM and glycaemic traits during pregnancy in an East Asian population and highlight the potential role of inflammatory pathways in the aetiology of GDM and variations in glycaemic levels.

**Data availability:** Full summary statistics of GDM, FPG, OGTT0H, OGTT1H and OGTT2H will be made available in the GVM database and the GWAS Catalog upon publication.

**Research in context:** *What is already known about this subject?:*

- Previous genome-wide association studies on gestational diabetes mellitus (GDM) and glycaemic levels in Asian populations have been limited to sample sizes of a few thousand participants.
- There is a lack of research investigating the potential causal relationships between various pregnancy indicators and GDM as well as glycaemic levels.

*What is the key question?:*

- What are the genetic and molecular factors that contribute to GDM and glycaemic levels in an under-represented population of Chinese pregnant women in East Asia?

*What are the new findings?:*

- Four genetic loci were significantly associated with GDM risk, with *MTNR1B* being the most prominent. A total of 31 genetic loci, including 14 novel loci, were significantly associated with four commonly measured glycaemic traits.
- A genetically higher lymphocyte percentage and lower white blood cell count, neutrophil percentage and absolute neutrophil count were associated with increased glucose levels and an elevated risk of GDM.

*How might this impact on clinical practice in the foreseeable future?:*

- Individuals with GDM risk variants could receive special attention during early pregnancy to mitigate the risk of adverse pregnancy outcomes associated with GDM and higher glycaemic levels. Clinical trials may be warranted to examine the potential of interventions such as nutrition therapy or medication for modulating inflammatory responses and reducing GDM incidence.

## Introduction

Gestational diabetes mellitus (GDM), defined as glucose intolerance resulting in hyperglycaemia that begins or is first diagnosed in pregnancy, has become the most prevalent medical disorder during pregnancy [1]. The global prevalence of GDM has significantly increased over the past few decades, affecting 16.7% of pregnant women worldwide, with an estimated 21 million livebirths affected in 2021 [2]. In mainland China, GDM prevalence among pregnant women is 14.8% (95% CI 12.8%, 16.7%), leading to substantial healthcare and economic burdens [3]. Recognised for its role in the ‘diabetes begetting diabetes’ circle, early diagnosis and management of GDM during pregnancy are considered crucial strategies to prevent adverse birth outcomes and reduce the long-term burden of chronic metabolic conditions such as type 2 diabetes [4, 5].

Genetic variation significantly influences susceptibility to human diseases, providing valuable insights into disease aetiology and innovative approaches to prevention and treatment [6]. However, our understanding of key genes and molecular pathways related to GDM onset remains limited because of the challenges in collecting extensive genetic data and conducting RCTs in pregnant populations. Current genome-wide association studies on GDM in Asian populations have been constrained to relatively small sample sizes, typically in the hundreds or thousands [7]. Unlike type 2 diabetes, for which twin studies have estimated heritability of 72% [8], the heritability of GDM remains unclear. Furthermore, while those with GDM demonstrate individual variability in disease severity, as reflected in glycaemic levels and medication requirements, there is a dearth of knowledge regarding the genetic and molecular factors contributing to this phenotypic spectrum.

In this study, we aimed to integrate data from a routine pregnancy screening programme in Shenzhen City, China, including genetic data obtained through non-invasive prenatal testing (NIPT) [9], blood test measurements and comprehensive electronic medical records, from a sample of 30,699 pregnant women to investigate the genetic and molecular risk factors associated with GDM. This is the largest genome-wide association study (GWAS) of GDM susceptibility in an Asian population to date, involving 3317 participants with GDM (14.5%) and 19,565 control participants without GDM (85.5%), and including four quantitative glycaemic traits (fasting plasma glucose [FPG] and 0 h, 1 h and 2 h OGTTs [OGTT0H, OGTT1H and OGTT2H, respectively]).

Additionally, we explored potential causal associations between 55 biochemical biomarkers assessed early in the first or second trimester during pregnancy screening and GDM and glycaemic levels in pregnancy using Mendelian randomisation (MR).

## Methods

### Study design and participants

NIPT, a genomic sequencing technology for identifying fetal trisomy by sequencing maternal plasma-free DNA (cfDNA), has been widely used in screening programmes in pregnancy since 2011 [10]. In our previous study, we established the utility of NIPT data for GWAS analysis [9]. In this study, we recruited 30,699 Chinese pregnant women who visited Shenzhen Baoan Women’s and Children’s Hospital (Shenzhen, China) for maternity check-ups and received an NIPT test in the first or second trimester between 2017 and 2019. The birth place distribution of participants is detailed in ESM Table 1, with a predominant concentration in southern China, representing southern Chinese. Maternal genotypes were inferred from low-depth whole-genome sequencing data generated from NIPT using previously described methods [9]. Blood tests were conducted to assess 55 biomarkers (ESM Table 2) and the FPG level during gestational weeks 16–18. The remaining three glycaemic traits (OGTT0H, 1H and 2H) were assessed during gestational weeks 24–28. Physical measurements, including maternal age, gestational week, maternal weight, early pregnancy BMI and blood pressure, as well as the diagnosis of GDM, were obtained through the hospital’s electronic medical record system. In the case of multiple records, we used the first measurements taken during gestational weeks 13–18. Among the 30,699 participants, 7817 participants did not have complete records for glucose tests and clinical diagnosis and were excluded from the GWAS analysis of GDM. Of the remaining participants, 3317 were diagnosed as having GDM and 19,565 were included as control participants.

**Table 1.**
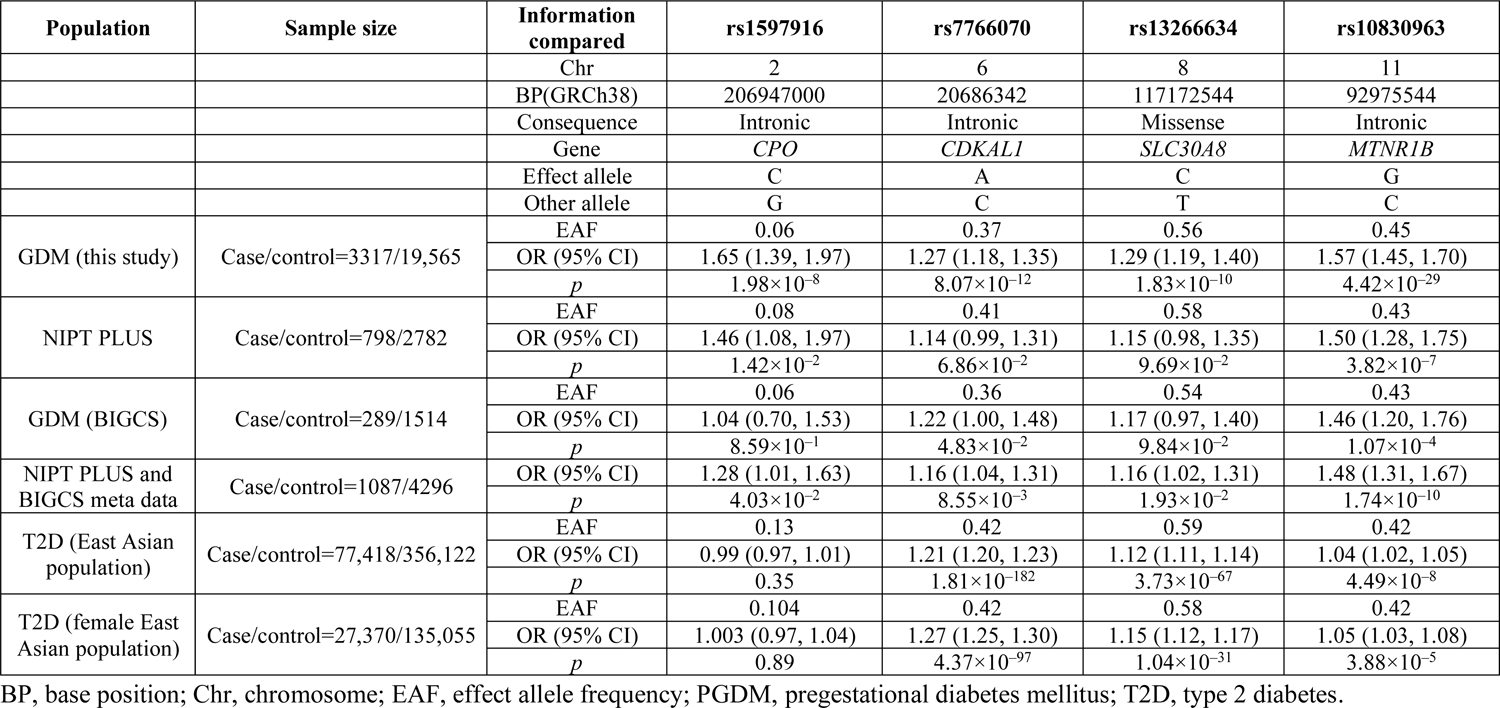
Comparison of genome-wide significant association signals.

We integrated the genetic data, blood test results and medical records to investigate the genetic and molecular risk factors underlying the phenotypic spectrum of GDM and glycaemic traits. The overall study design is shown in Fig. 1.This study received approval from the Institutional Review Board of the School of Public Health (Shenzhen), Sun Yat-sen University (2021.No.8), and the Institutional Board of Shenzhen Baoan Women’s and Children’s Hospital (LLSC2019-07-11-KCW). All study participants provided written informed consent.

**Fig. 1.**
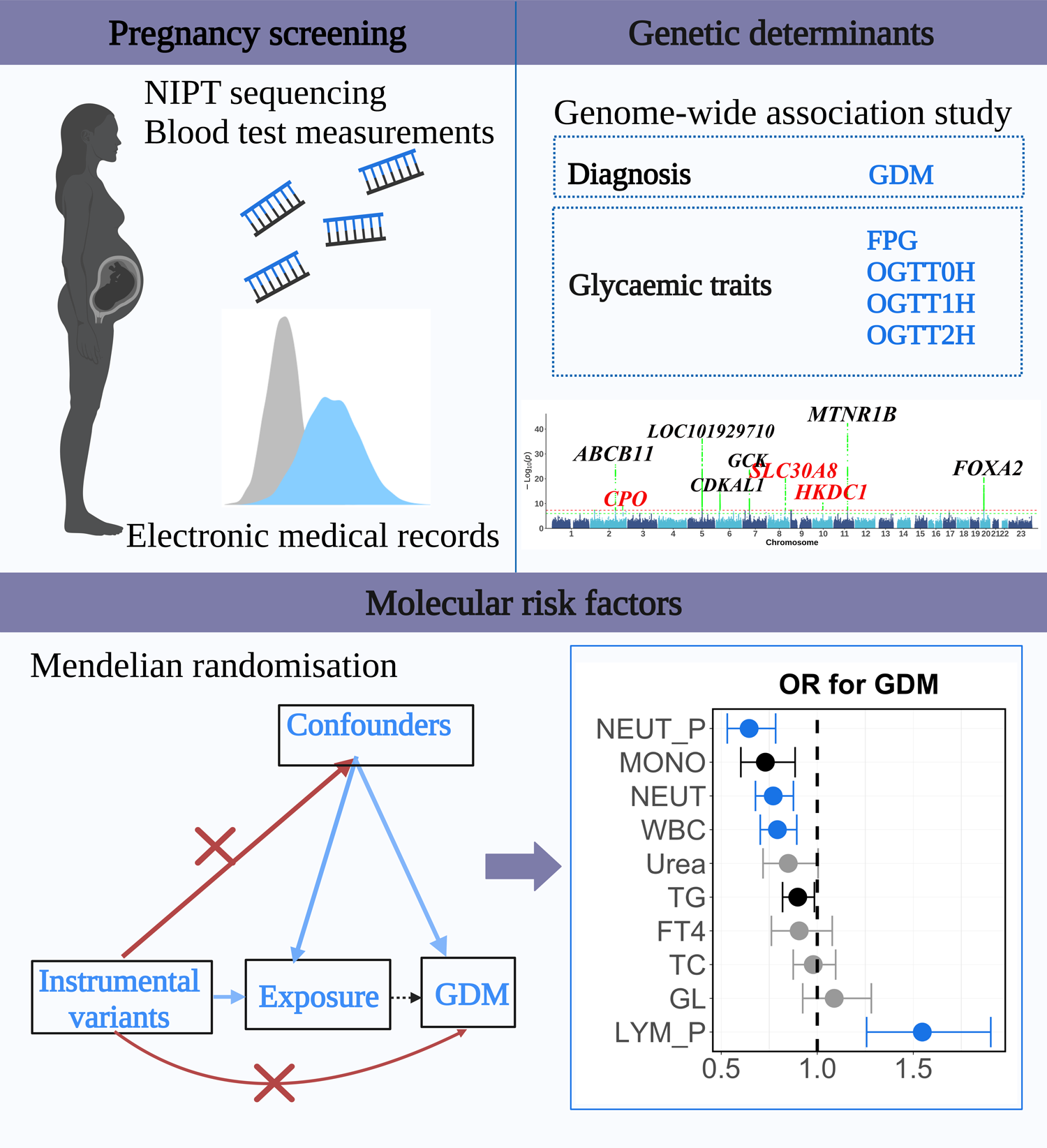
Study design, depicting the data sources and analyses performed. We collected NIPT sequencing data, blood test data and information from electronic medical records. Subsequently, a GWAS was conducted to investigate the associations between NIPT-derived genotypes and GDM diagnosis, baseline glycaemic levels and glycaemic levels post-glucose challenge. Additionally, one-sample MR analyses to explore potential causal relationships between the biochemical measurements and the risk of GDM and glycaemic levels were carried out. FT4, free thyroxine; GL, globulin; LYM_P, lymphocyte percentage; MONO, absolute monocyte count; NEUT, absolute neutrophil count; NEUT_P, neutrophil percentage; TC, total cholesterol; TG, triglyceride; WBC, white blood cell count. Created with BioRender.com

### Phenotype definition

After excluding participants with pregestational diabetes mellitus, GDM was diagnosed by administering a 75 g OGTT during gestational weeks 24-28. A participant was diagnosed with GDM if any of the following criteria were met: (1) OGTT0H glucose level ≥5.1 mmol/l; (2) OGTT1H glucose level ≥10.0 mmol/l; or (3) OGTT2H glucose level ≥8.5 mmol/l [11]. Glucose levels were measured using a Beckman Coulter (America) AU(GLU) kit and assayed using an AU5800 biochemical analyser.

### Statistical analyses

Means and SDs were used to depict the basic characteristics of the study participants (ESM Table 3). The associations of GDM and the four quantitative glycaemic traits (FPG, OGTT0H, OGTT1H and OGTT2H) with the 55 biomarkers were assessed using multiple regression analyses, in which maternal age, maternal BMI and the gestational week of the OGTT tests were included as covariates. To facilitate the comparison of results, the quantitative dependent and independent variables in the regression analyses were normalised using quantile transformation. All statistical analyses were conducted using R (version 4.0.4, https://mirror-hk.koddos.net/CRAN/, accession date: 4/1/2023).

### Genotype imputation from NIPT data

Genotype imputation was performed using STITCH software (version 1.2.7, http://www.well.ox.ac.uk/rwdavies/stitch_2017_02_14.html, accession date: 15/2/2023), as described in our previous study [9]. Following imputation, we selected variants with an information score >0.4, a minor allele frequency (MAF) <0.01 and a Hardy–Weinberg equilibrium *p* value <10^−6^ for subsequent analysis [9]. The average imputation accuracy, evaluated in a subset of 110 participants with high-coverage sequencing data (>20×), was 0.89, consistent with previous assessments [9].

### Genome-wide association analysis

Following rigorous quality control of the sequencing data, including principal component analysis and genotype imputation using an established protocol [9], we conducted GWAS to examine the association of GDM, the four quantitative glycaemic traits (FPG, OGTT0H, OGTT1H and OGTT2H) and the 55 biomarkers with the imputed genotype dosage from NIPT, assuming additive genotype effects. The GWAS was conducted using a multiple linear regression or logistic regression model implemented in PLINK 2.0 [12]. We included gestational week, maternal age, maternal BMI and the top five principal components to account for population stratification as covariates. GWAS summary statistics, including effect size, SE and *p* value, were used for subsequent genome-wide and regional association plots and MR analyses. We compiled the Manhattan and quantile–quantile (QQ) plots using an R (version 4.0.4) script and generated regional association plots using Locuszoom (version 1.4) [13].

To replicate GWAS findings for GDM, FPG, OGTT0H, OGTT1H and OGTT2H, we employed two additional cohorts: the NIPT PLUS cohort, comprising 4688 independent pregnancies with deeper sequencing of maternal plasma-free DNA (ESM Methods: ‘The NIPT PLUS cohort’), and the Born in Guangzhou Cohort Study (BIGCS) [14], encompassing whole-genome sequencing and electronic medical records for 1,854 unrelated pregnancies. As well as individual replication within these two independent cohorts, we conducted a meta-analysis using METAL (release 25 March 2011, http://www.sph.umich.edu/csg/abecasis/Metal/, accession date: 15/9/2023) and replicated our findings with the results of this meta-analysis. Differences in SNP genetic effects between our study and the meta-analysis of the two independent cohorts were assessed using a heterogeneity test integrated into METAL, along with a comparison of the direction and significance of effect size estimates. Furthermore, we compared the

GWAS results for FPG and OGTT2H with summary statistics data from the MAGIC consortium (https://www.magicinvestigators.org/, accession date: 4/3/2023) [15]. Additionally, we applied a two-sample *t* test to examine the different genetic effects on baseline glucose levels and glucose levels after an OGTT (ESM Methods: ‘Statistical test of different genetic effects between two GWAS’).

### Genetic correlation

We employed linkage disequilibrium (LD) score regression [16] to assess genetic correlations between GDM and glycaemic traits in our study and type 2 diabetes in an East Asian population comprising 77,418 type 2 diabetes cases and 356,122 controls [17] and a European population comprising 74,124 type 2 diabetes cases and 824,006 controls [18]. We also estimated genetic correlations in the female subset of the East Asian population, including 27,370 type 2 diabetes cases and 135,055 controls [17]. These genetic correlations were estimated using the same LD score regression settings employed in heritability calculations [19].

### Mendelian randomisation analyses

We employed bidirectional MR analyses using two-sample methods within a single cohort [20] and the TwoSampleMR package [21] to identify potential causal associations between biomarkers and both GDM and glycaemic levels. We used genome-wide complex trait analysis–conditional and joint association analysis (GCTA-COJO) and selected independent SNPs with a *p* value <5×10^-8^ as genetic instruments [22], using high-depth sequencing data consisting of 10,000 Chinese people as a reference panel [23]. In this analysis, we excluded loci with a MAF <0.01, and the threshold of the LD *r*^2^ was set at 0.2 (command of analysis: gcta64 –bfile reference_panel_file –cojo-file gwas_file –cojo—slct –cojo-p 5e-8 -- cojo-collinear 0.2 --maf 0.01). We restricted valid genetic instruments to those with *F* statistics >10.

To test the independence and restriction exclusion assumptions for MR, we conducted pleiotropy and heterogeneity tests. We also performed sensitivity analyses using alternative MR methods, including MR Egger, simple mode, weighted median and weighted mode. Among these methods, we primarily relied on the inverse variance weighted (IVW) method, known for its superior statistical power in estimating potential causal associations [24]. We considered potential causal estimates as significant when they met a Bonferroni-adjusted *p* value threshold in IVW analysis and either showed no evidence of heterogeneity (*p*≥0.05) and horizontal pleiotropy (*p*≥0.05) or, in the presence of heterogeneity or horizontal pleiotropy (*p*<0.05), a least one of the additional estimates from MR Egger, simple mode, weighted median or weighted mode was significant (*p*<0.05). To calculate the power of the MR analyses, we used an online tool (http://cnsgenomics.com/shiny/mRnd/, accessed 20 September 2023) [25].

To ensure the reliability of the MR results, we conducted a meta-analysis using METAL (release 25 March 2011) between this study and NIPT PLUS. Subsequently, we performed bidirectional MR analyses on the metadata [21] using the TwoSampleMR package [22].

## Results

### Clinical and epidemiological characteristics of the study participants

We recruited 30,699 eligible pregnant women between 2017 and 2019. After excluding 163 individuals with pregestational diabetes based on clinical diagnoses, 3317 women (10.8%) were diagnosed with GDM and 19,565 were included as control participants without diabetes (ESM Table 3). Age, weight, BMI and blood pressure exhibited significant associations with both GDM and glycaemic levels (ESM Table 4).

We employed multiple regression analysis to examine potential associations between specific biomarkers, measured during early pregnancy screening (16–18 weeks of gestation), and GDM, as well as four quantitative glycaemic traits measured during pregnancy. GDM and the four glycaemic traits (FPG, OGTT0H, OGTT1H and OGTT2H) were regressed on 55 pregnancy screening biomarkers categorised into seven biological categories—routine blood tests (*n*=28), biomarkers for anaemia (*n*=6), blood lipids (*n*=2), kidney function (*n*=3), liver function (*n*=8), thyroid function (*n*=3) and infection (*n*=5) (ESM Table 2)—adjusting for BMI, maternal age and gestational week. Following Bonferroni correction, we found positive associations beween the onset of GDM and triglyceride, 25-hydroxyvitamin D and folic acid levels, haematocrit and red blood cell count, while total bile acid and triglyceride levels exhibited positive associations with FPG and OGTT2H, respectively (ESM Fig. 1). Conversely, total bilirubin level, immature granulocyte count and percentage and monocyte absolute value and percentage showed negative associations with GDM, and monocyte percentage showed a negative association with FPG (ESM Fig. 1).

### Genetic determinants of GDM and glycaemic traits among Chinese pregnant women

To investigate the genetic factors underlying GDM susceptibility and the distribution of the four glycaemic traits, we conducted the following GWAS analyses: (1) GDM cases (*n*=3317) vs controls (*n*=19,565); and (2) linear regression on FPG (*n*=26,751), OGTT0H (*n*=24,929), OGTT1H (*n*=24,931) and OGTT2H (*n*=24,931). Our power analysis indicated that we had the ability to identify genetic loci with a minimum MAF of 0.01 and a minimum OR of 1.4, or a minimum MAF of 0.05 and a minimum OR of 1.2, or a minimum MAF of 0.2 and a minimum OR of 1.1 for GDM analysis. For the analysis of the four glycaemic traits, we could detect loci with a minimum MAF of 0.01 and an effect size of 0.29 (ESM Fig. 2). We estimated the SNP heritability (*h^2^_g_*) for GDM to be 3.2% (s.e. 1.9%) and for the four glycaemic traits to be between 5.3% and 10.2% (s.e. 2.2%) (ESM Table 5). We found a strong genetic correlation between GDM and type 2 diabetes in the female subset of the East Asian population from Spracklen et al [16] (*r_g_*=0.84, s.e.=0.31, *p*=0.006), as well as between GDM and the total East Asian population (*r_g_*=0.81, s.e.=0.24, *p*=8.0×10^-4^), but not between GDM and the European population from Mahajan et al [17] (*r_g_*=0.51, s.e.=0.21, *p*=0.013) (ESM Table 5). In addition, our study, in conjunction with data from the MAGIC consortium [15], suggests higher heritability for OGTT2H than FPG (ESM Tables 5 and 6).

**Fig. 2.**
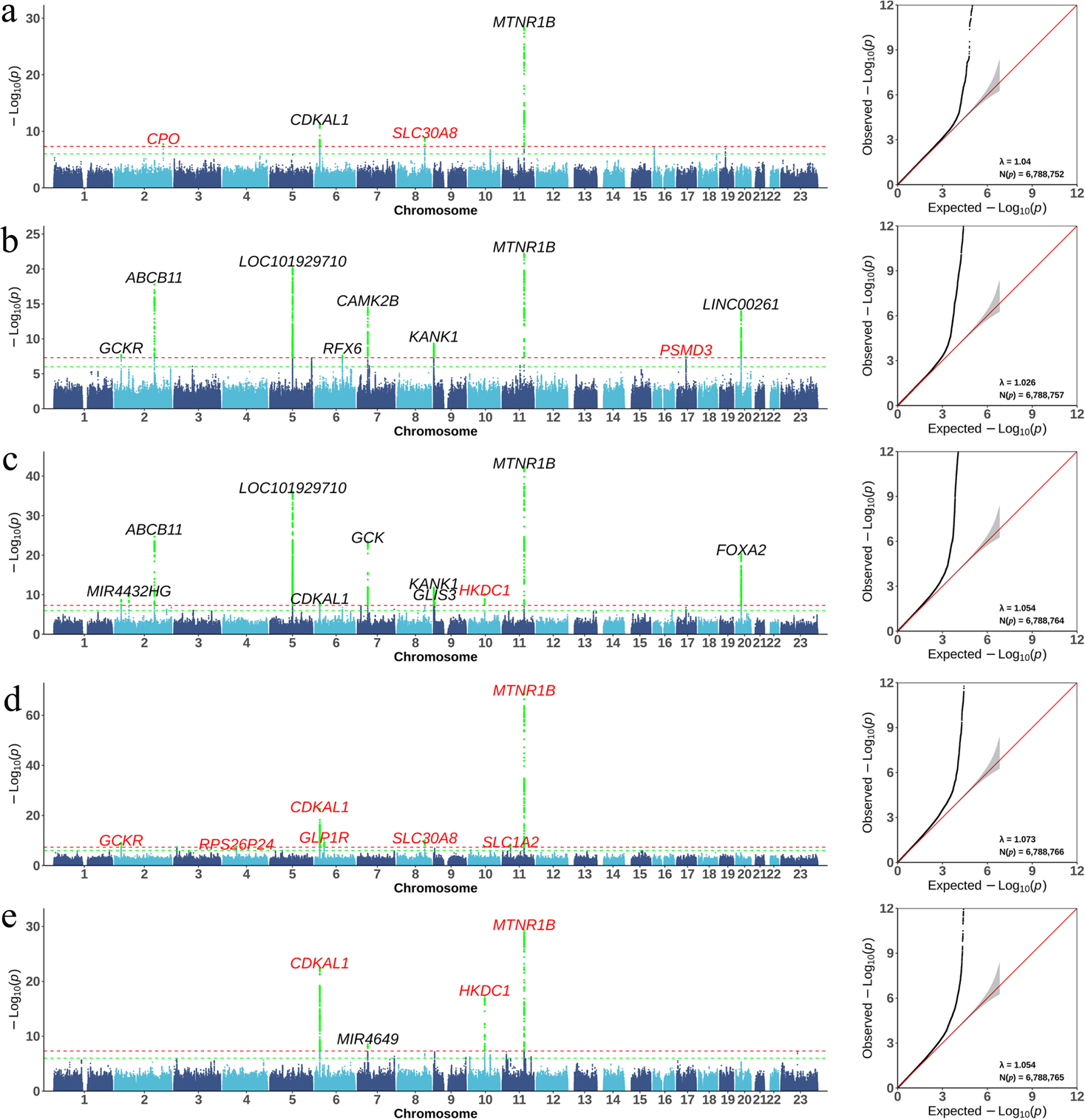
Manhattan and QQ plots for GDM and the four glycaemic traits in pregnancy. GWAS results for (**a**) GDM, (**b**) FPG, (**c**) OGTT0H, (**d**) OGTT1H and (**e**) OGTT2H. In the Manhattan plots, horizontal lines define genome-wide significance (*p*=5×10^−8^; red) and suggestive significance (*p*=1×10^−6^; green) thresholds. Newly identified loci are denoted in red, while loci with established knowledge are denoted in black (genome-wide association study *p* values <5×10^−8^ and that were previously reported in the GWAS Catalog version 1.0.2).

We identified four genome-wide significant loci associated with GDM susceptibility. The most significant locus was situated within the intron of the *MTNR1B* gene (lead SNP rs10830963-G, OR [95% CI] 1.57 [1.45, 1.70], *p*=4.42×10^-29^), while three additional loci were identified within the *CDKAL1* gene (lead SNP rs7766070-A, intronic variant, OR [95% CI] 1.27 [1.18, 1.35], *p*=8.07×10^-12^), *SLC30A8* gene (lead SNP rs13266634-C, missense variant, OR [95% CI] 1.29 [1.19, 1.40], *p*=1.83×10^-10^) and *CPO* gene (lead SNP rs1597916-C, intronic variant, OR [95% CI] 1.65 [1.39, 1.97], *p*=1.98×10^-8^) (Fig. 2a, Table 1).

All four loci were replicated in the meta-analysis of two independent cohorts: the BIGCS birth cohort [14], comprising 289 GDM cases and 1514 controls, and the NIPT PLUS cohort, comprising 798 GDM cases and 2782 controls. These findings showed consistent effect directions and no statistically significant differences in the effect estimates between our study and the meta-analysis of the independent cohorts (*p_het_*>0.05) (ESM Table 7). Notably, while rs10830963 exhibits the most pronounced genetic effect on GDM (OR [95% CI] 1.57 [1.45, 1.70], *p*=4.42×10^-29^), it does not serve as a major genetic determinant for female type 2 diabetes (OR [95% CI] 1.05 [1.03, 1.08], *p*=3.88×10^-5^) or all type 2 diabetes (OR [95% CI] 1.04 [1.02, 1.05], *p*=4.49×10^-8^) in the studied East Asian population [16] (Table 1). This indicates a different genetic aetiology between GDM and type 2 diabetes, despite their high genetic correlation. Additionally, in a GWAS analysis of all GDM cases (*n*=3317) when comparing those receiving insulin (*n*=419) with those not receiving glucose-lowering medication (*n*=2898), no significant loci were identified (ESM Fig. 3). While further replication and validation are necessary, these findings suggest that genetics may have a limited role in explaining susceptibility to insulin medication in people with GDM.

**Fig. 3.**
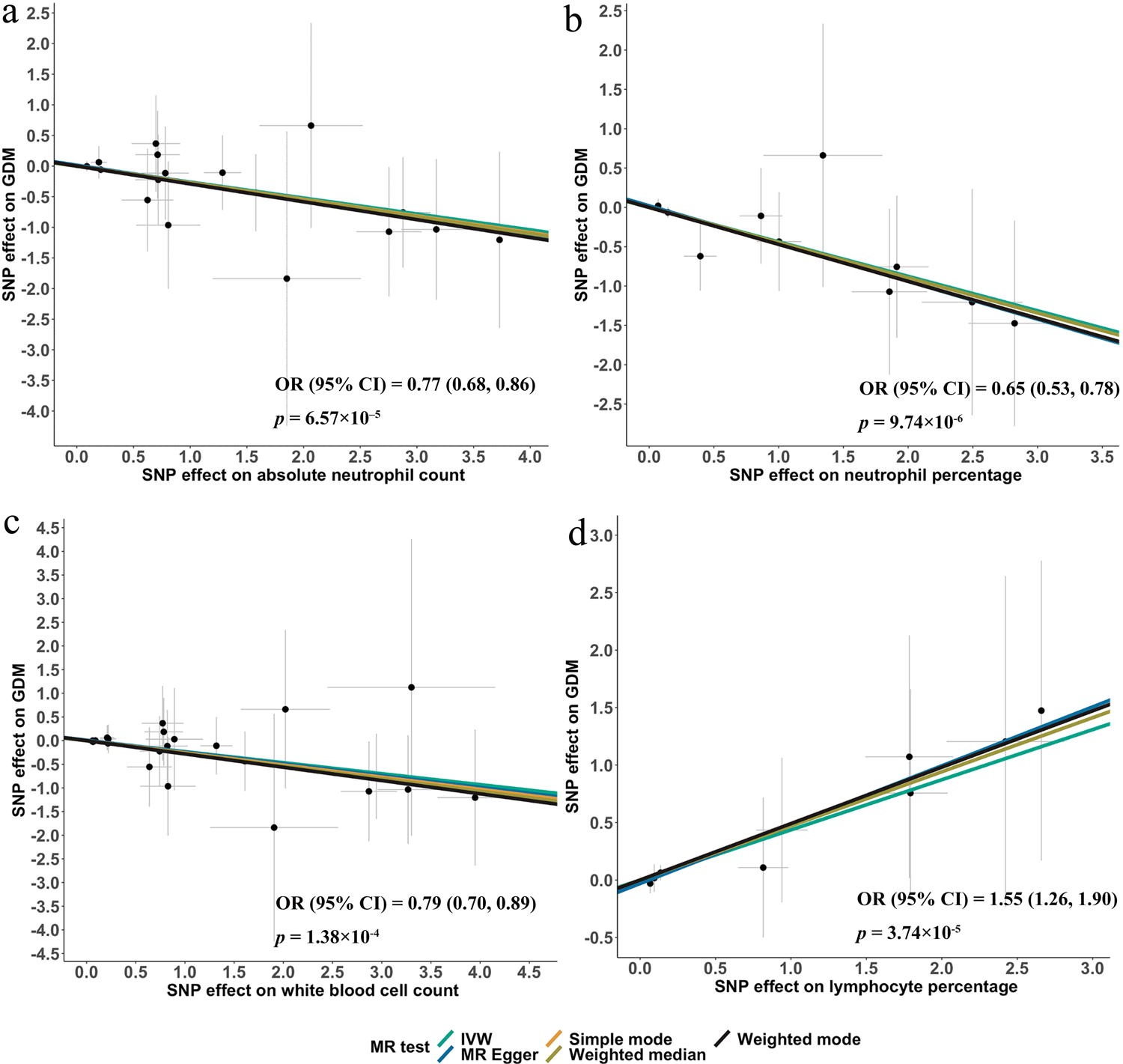
Scatterplots of MR analysis results with biomarkers as the exposures and GDM as the outcome: (**a**) Absolute neutrophil count, (**b**) neutrophil percentage, (**c**) white blood cell count and (**d**) lymphocyte percentage. The effect size and *p* values for the IVW analysis are shown in the figure. The slope of the regression line in each panel indicates the direction of the effect of the exposure on GDM, with a positive slope representing a positive effect and a negative slope indicating a negative effect. The effects of these biomarkers on the four glycaemic traits are shown in ESM Fig. 8

We also identified 31 loci that were significantly associated with the four glycaemic traits (*p*<5×10^-8^), including 14 newly discovered associations (Fig. 2b–e, ESM Table 8). These 31 loci were compared with GWAS data from the meta-analysis of the NIPT PLUS and BIGCS cohorts for the four glycaemic traits, and the NIPT PLUS cohort for FPG. The replication rates for FPG, OGTT0H, OGTT1H and OGTT2H were six out of nine, 10 out of 11 five out of seven and four out of four, respectively, showing a consistent direction of effect and a lack of heterogeneity between our study and the independent cohorts (ESM Table 7). Notably, the consistency rates were six out of nine and one out of four for the nine and four significant associations for FPG and OGTT2H, respectively, using the East Asian summary statistics from the MAGIC consortium (ESM Table 9). A comparison of the genome-wide association analysis for FPG and OGTT2H between the MAGIC East Asian population and our study suggests a lack of power for OGTT2H in MAGIC and a potential genetic difference between the East Asian population included in MAGIC and the Chinese population included in this study (ESM Fig. 4). This analysis included several notable discoveries. First, *MTNR1B* was the most significant signal for all of the glycaemic traits, including OGTT1H, which was not previously investigated, and OGTT2H, for which no *MTNR1B* association was previously reported in either the East Asian or European populations [15]. Second, the genetic determinants of FPG (examined in the first trimester) and OGTT0H (examined between 24 and 28 gestational weeks) were highly similar, sharing seven significant genetic loci (Fig. 2b, c). Third, despite a similar sample size, the genetic determinants of the baseline glycaemic level (i.e. FPG and OGTT0H) demonstrated substantial differences from the genetic determinants of the glycaemic level after a glucose challenge (i.e. OGTT1H and OGTT2H) (Fig. 2d, e).

We compared the genetic effects of the FPG lead SNPs on OGTT1H and OGTT2H, revealing an inconsistency rate of seven out of nine and eight out of nine for OGTT1H and OGTT2H, respectively (ESM Table 10), reflecting the genetic differences in fasting glucose levels and glucose levels in response to an OGTT. After adjusting for FPG, the principal effector genes associated with OGTT1H and OGTT2H remained unaltered, implying that these genes are involved in glucose regulation and remain unaffected by variations in fasting glucose levels. It is noteworthy that *MTNR1B* influences FPG, OGTT1H and OGTT2H at the same time. In addition, *CDKAL1* exerts an influence on both OGTT1H and OGTT2H glucose levels, while having no impact on FPG. Furthermore, *HKDC1* affects OGTT2H glucose levels without influencing FPG or OGTT1H (ESM Fig. 5). Specifically, four loci, including the lncRNA gene locus *LOC101929710* (lead SNP rs10476553), previously known to be associated with BMI, the *ABCB11* (lead SNP rs853774) locus, related to severe cholestatic liver disease, the *GCK* (lead SNP rs730497) locus, associated with multiple types of diabetes, and the *FOXA2* (lead SNP rs6048209) locus, linked to MODY, were specifically associated with baseline FPG and OGTT0H glucose levels but not OGTT1H and OGTT2H glucose levels (Fig. 2c-e, ESM Table 8).

Finally, there were several loci that were significantly associated with the glycaemic traits (*p*<5×10^-8^) but that did not contribute to GDM susceptibility, such as the abovementioned *LOC101929710*, *ABCB11*, *GCK* and *FOXA2* loci. The regional association plots showing all of the genes in a 1 Mbp window flanking the lead SNP generated by Locuszoom are presented in ESM Fig. 6.

### Potential causal effects of 55 biomarkers on GDM and glycaemic levels

In our study, we found that genes involved in circadian rhythm regulation and glucose homeostasis have a significant effect on GDM susceptibility and glycaemic levels. We also investigated whether specific biochemical measurements (*n*=55; ESM Table 2, reflecting various body functions and assayed during early pregnancy (16–18 weeks of gestation) as part of the pregnancy screening programme, may have a potential causal effect on GDM or glycaemic levels. To achieve this, we employed bidirectional MR analysis using a two-sample method on a single dataset [20]. This approach underscores the reliability of applying two-sample MR methods for one-sample MR when sufficient instrumental variable strength is present (see Methods).

Based on the IVW MR model results, we identified several significant potential causal associations between routine blood biomarkers and GDM susceptibility (Fig. 3, ESM Table 11) as well as glycaemic levels in pregnancy (ESM Figs 7 and 8). Genetically higher absolute neutrophil count (OR [95% CI] 0.77 [0.68, 0.86], *p*=6.57×10^-5^), neutrophil percentage (OR [95% CI] 0.65 [0.53, 0.78], *p*=9.47×10^-6^) and total white blood cell count (OR [95% CI] 0.79 [0.70, 0.89], *p*=1.38×10^-4^) demonstrated significant protective effects on GDM risk (Fig. 3a–c) and resulted in decreased FPG and OGTT0H glucose levels (ESM Figs 7 and 8). Conversely, a genetically higher lymphocyte percentage was associated with increased GDM risk (OR [95% CI] 1.55 [1.26, 1.90] *p*=3.74×10^-5^) (Fig. 3d) and elevated FPG and OGTT0H glucose levels (ESM Figs 7 and 8). All instrumental variables for these biomarkers are presented in ESM Table 12, and they exhibited no heterogeneity or pleiotropy (ESM Table 11). Additionally, in the MR analyses with GDM as the outcome, the statistical power exceeded 0.98 for absolute neutrophil count, lymphocyte percentage, neutrophil percentage and white blood cell count as exposures, while the *F* statistics for these four measurements exceeded 1000 (ESM Table 13). These results were consistent with the direction of the effect in the observational associations (ESM Fig. 1). The reverse MR analysis revealed no potential causal association of GDM with the aforementioned biomarkers (ESM Figs 9 and 10).

Furthermore, we conducted bidirectional MR analyses using meta-analysis results from the discovery study and the NIPT PLUS cohort to investigate the associations between absolute neutrophil count, lymphocyte percentage, neutrophil percentage, white blood cell count and GDM, as well as glycaemic levels. These findings closely aligned with the MR analyses conducted without the NIPT PLUS cohort. Specifically, a genetically higher total white blood cell count (OR [95% CI] 0.81 [0.72 0.92], *p*=8.97×10^-4^), absolute neutrophil count (OR [95% CI] 0.79 [0.69 0.90], *p*=4.44×10^-4^) and neutrophil percentage (OR [95% CI] 0.72 [0.59, 0.91], *p*=5.06×10^-3^) showed significant protective effects on GDM risk (ESM Fig. 11a-c) and resulted in decreased FPG and OGTT0H glucose levels (ESM Table 14). A genetically higher lymphocyte percentage was significantly associated with lower FPG and OGTT0H glucose levels, although the potential causal relationship between lymphocyte percentage and GDM showed less significance.

## Discussion

In this study, we harnessed extensive genetic and phenotypic data from a standard pregnancy screening programme to explore the genetic and molecular factors associated with GDM susceptibility and glycaemic levels during pregnancy. We identified four significant loci (*MTNR1B*, *CDKAL1*, *SLC30A8* and *CPO*) associated with GDM susceptibility at a genome-wide level. The most significant locus, *MTNR1B*, encodes the melatonin receptor 1B, predominantly found in the retina and brain, where it plays a role in light-dependent functions and regulates the reproductive and circadian effects of melatonin. This receptor also exerts an inhibitory influence on insulin secretion.[26] and previous studies have linked the lead SNP (rs10830963) located in the intron of the *MTNR1B* gene to FPG levels, HbA_1c_ measurements, type 2 diabetes and birth weight [7].

While a Korean study of GDM [27] with a smaller sample size (1399 cases and 2025 controls) previously reported both *MTNR1B* and *CDKAL1* as GDM-associated loci, with *CDKAL1* exhibiting the largest genetic effect, our study, along with the NIPT PLUS and BIGCS replication cohorts, indicates that *MTNR1B* plays a more substantial role in GDM susceptibility than *CDKAL1* (Table 1). Importantly, our study rigorously excluded participants with pregestational diabetes from the GDM group, providing a more accurate picture of the genetic determinants of GDM susceptibility than the Korean study. Furthermore, our study, with nearly ten times the sample size of the Korean study, enables more precise estimation of the genetic effect size. These two loci also have varying roles in the risk of developing type 2 diabetes. For type 2 diabetes in both East Asian and European populations, *CDKAL1* was recognised as the second strongest genetic locus [17, 28], whereas the *MTNR1B* locus either barely passed or did not reach the genome-wide significance threshold (Table 1), underscoring the differences in genetic architecture between GDM and type 2 diabetes.

Furthermore, we identified 31 genetic loci associated with four glycaemic traits (FPG, OGTT0H, OGTT1H and OGTT2H) that are commonly used for GDM diagnosis. Fourteen of these loci were newly discovered associations. The high replication rate of the FPG loci and better performance of the OGTT2H loci when compared with the MAGIC consortium indicates the reliability of our discoveries and supports the value of using low-pass NIPT sequencing data to investigate genotype–phenotype correlations [9]. In the GWAS of the four glycaemic traits, the *MTNR1B* locus, which was previously known to be associated with FPG [7], was first discovered to have the greatest genetic effect on OGTT1H and OGTT2H. Notably, we identified substantial differences in the genetic determinants of baseline glycaemic levels (FPG and OGTT0H) and glycaemic levels after an oral glucose challenge (OGTT1H and OGTT2H), which mainly involved four genetic loci in *LOC101929710*, *ABCB11*, *GCK* and *FOXA2.* These four genetic loci specifically contribute to FPG and OGTT0H but not OGTT1H and OGTT2H, indicating that there are distinct biological mechanisms regulating the baseline glucose concentration and glucose metabolism after glucose intake. Furthermore, many of the genetic loci that play a major role in regulating glycaemic concentrations, such as the abovementioned *LOC101929710*, *ABCB11*, *GCK* and *FOXA2* loci, do not contribute to GDM susceptibility.

Of the 55 biochemical measurements, which were categorised into seven groups (routine blood biomarkers, biomarkers of anaemia, blood lipid, kidney function, liver function, thyroid function and infection), four immune cell measurements (white blood cell count, lymphocyte percentage, absolute neutrophil count and neutrophil percentage) in the routine blood biomarkers category demonstrated potential causal risk or protective effects on GDM and glycaemic levels (Fig. 3). Previous research has hinted at associations between obesity and type 2 diabetes and the accumulation of immune cells, including lymphocytes and neutrophils and other subtypes of immune cells, in various tissues [29]. However, establishing a potential causal link between inflammation and metabolic dysfunction remains challenging. Therefore, we conducted a bidirectional MR analysis, which suggested that inflammation may potentially contribute to GDM risk and higher FPG and OGTT0H glucose levels. Notably, this potential effect on GDM aligns with its impact on FPG and OGTT0H glucose levels but not OGTT1H and OGTT2H glucose levels after a glucose challenge.

It is important to note the limitations of our study and potential directions for future research. Despite being the largest GWAS on GDM in Asia to date, our design was limited to common variants (MAF>0.01) with intermediate or mild effects. Investigating low-frequency and rare variants may uncover additional genetic risks associated with GDM [30]. Furthermore, although our study provides new insights into the potential causal role of circadian rhythm regulation, glucose homeostasis, inflammation and oxidative stress pathways as genetic and molecular risk factors for GDM, functional validation and clinical trials are essential for a deeper understanding of these findings.

We must also acknowledge that instrumental variable analysis focuses on estimating the effects of fixed-point or time-stable exposures, which might not account for ‘lifetime effects’ [23] Assessing temporal variations in exposures was challenging because of the lack of pre-pregnancy measurements for these 55 biomarkers. Although we managed to replicate the MR outcomes using available metadata, potential bias in MR estimates might exist if the exposures demonstrated substantial lifetime variability. Future research should consider functional validation to confirm these findings.

Finally, it is noteworthy that factors such as family history of diabetes, educational background and lifestyle factors may influence GDM susceptibility. However, as these factors do not influence an individual’s genotype and were not considered confounding factors, their impact on the GWAS and MR results in this study is likely to be minimal. Nevertheless, it will be valuable to explore how adjusting for these factors affects the study’s conclusions in future research studies.

Data availability Full summary statistics of GDM, FPG, OGTT0H, OGTT1H and OGTT2H will be made available in the GVM database and the GWAS Catalog upon publication.

## Authors’ relationships and activities

The authors declare that there are no relationships or activities that might bias, or be perceived to bias, their work.

## Contribution statement

SL and LX designed the study. JZ, WW, HL, YY and QC collected and organised data from the maternity testing system. JZ, YG, PW and SL conducted data pre-processing and the preliminary analyses. YG, PW, SB and SL performed all regression analyses, GWAS and MR. YG and PW performed the visualisation of all results. SH, MH and XQ provided the validation data. SL, GJ, XQ and LX provided professional guidance and interpretation of data. YG, SL and JZ wrote the manuscript with input from all authors. All authors contributed to manuscript revisions and approved the final version of the article. SL is responsible for the integrity of the work as a whole.

## Abbreviations

BIGCS: Born in Guangzhou Cohort Study

FPG: Fasting plasma glucose

GDM: Gestational diabetes mellitus

GWAS: Genome-wide association study

IVW: Inverse variance weighted

LD: Linkage disequilibrium

MAF: Minor allele frequency

NIPT: Non-invasive prenatal testing

OGTT0H: 0 h OGTT

OGTT1H: 1 h OGTT

OGTT2H: 2 h OGTT

QQ: Quantile–quantile plot

## Data Availability

All data produced in the present study are available upon reasonable request to the authors

