## Supplemental material for "Genome-wide association and Mendelian randomisation analysis among 30,699 Chinese pregnant women identifies novel genetic and molecular risk factors for gestational diabetes and glycaemic traits"

**Electronic supplementary material(ESM) for**

Received: 11 August 2023 / Accepted: 3 November 2023

### ESM Methods

#### The NIPT PLUS cohort

In addition to the previously mentioned participants, we collected data from 4,688 individuals who sought maternity check-ups at Shenzhen Baoan Women's and Children's Hospital (Shenzhen, China) throughout the entire 40-week gestational period. These individuals underwent NIPT in either the first or second trimester between 2020 and 2021. Notably, these samples are characterized by a deeper sequencing depth in comparison to conventional NIPT, with an average depth of 0.3x. For this study, we leveraged their recorded data on physical measurement, blood glucose levels, lymphocyte percentage, white blood cell count, neutrophil percentage, absolute neutrophil count, and clinical diagnostic information related to GDM during pregnancy.

#### Statistical test of different genetic effects between two GWAS

In this study, we compared the GWAS results with those from MAGIC and between the baseline glucose and the challenged glucose levels. We examined the difference in genetic effects between two GWAS with a two-sided *two-sample t-test* with the following hypotheses:

$$Null\ hypothesis\ H_0: \beta_1 = \beta_2$$
$$Alternative\ hypothesis\ H_1: \beta_1 \neq \beta_2$$

The *T* statistic was computed as follows:

$$T = \frac{\beta_1 - \beta_2}{\sqrt{\frac{S_1^2}{n_1} + \frac{S_2^2}{n_2}}} = \frac{\beta_1 - \beta_2}{\sqrt{SE_1^2 + SE_2^2}} \sim t(v') \quad (1)$$

The degrees of freedom $v'$ was determined by the formula:

$$v' = \frac{(SE_1^2 + SE_2^2)^2}{\frac{SE_1^4}{n_1 - 1} + \frac{SE_2^4}{n_2 - 1}} \quad (2)$$

Herein, $\beta_1$ and $\beta_2$ represent the genetic effects associated with specific traits (for example, baseline glucose levels) and other traits (for example, challgend glucose levels), respectively. $S_1^2$ and $S_2^2$ denote sample variance, whereas $SE_1$ and $SE_2$ stand for estimated standard errors. It is established that the *T* statistic in equation (1) follows a *t*-distribution with a degree of freedom $v'$. To address potential inequality between $S_1^2$ and $S_2^2$ , the adjusted $v'$ was employed, computed according to formula (2).

For all the GWAS loci we conducted, we used the $P$-value threshold ($P < 0.05$) to define statistical significance and reported the specific genetic locus.

**Supplementary Tables**

**ESM Table 1 Geographic distribution of the participants in this study.**

| Province | Administrative Definiation of Region | Samplesize |
|---|---|---|
| Guangdong | South | 11161 |
| Hunan | South | 4911 |
| Hubei | Central | 3337 |
| Jiangxi | South | 2684 |
| Guangxi | South | 1909 |
| Henan | Central | 1303 |
| Sichuan | Central | 1193 |
| Anhui | Central | 511 |
| Shaanxi | North | 461 |
| Fujian | South | 450 |
| Shandong | North | 325 |
| Heilongjiang | North | 319 |
| Guizhou | South | 287 |
| Chongqing | Central | 286 |
| Jilin | North | 195 |
| Hebei | North | 170 |
| Liaoning | North | 161 |
| Hainan | South | 160 |
| Gansu | North | 154 |
| Jiangsu | Central | 152 |
| Shanxi | North | 130 |
| Yunnan | South | 122 |
| Zhejiang | Central | 114 |
| Neimenggu | North | 83 |
| Xinjiang | North | 38 |

| | | |
|---|---|---|
| Ningxia | North | 17 |
| Qinghai | North | 14 |
| Tianjin | North | 13 |
| Beijing | North | 3 |
| Shanghai | Central | 2 |
| Tibet | Central | 1 |

**ESM Table 2 55 biomarkers list that included in this study**

| Category | Biomarkers | Category | Biomarkers |
|---|---|---|---|
| Blood routine (N=28) | Absolute_neutrophils | Blood anemia (N=6) | 25_hydroxyvitamin_D_D2+D3 |
| | Absolute_value_of_basophils | | Ferritin |
| | Basophil_percentage | | Folic_acid |
| | Eosinophil_absolute_value | | HemoglobinA2 |
| | Eosinophil_percentage | | HemoglobinF |
| | Hematocrit | | Vitamin_B12 |
| | Hemoglobin_concentration | Blood lipid (N=2) | Total_cholesterolt |
| | Immature_granulocyte_percentage | | Triglyceride |
| | Immature_granulocytes | Kidney function (N=3) | Creatinine |
| | Large_platelet_ratio | | Urea |
| | Lymphocyte_absolute_value | | Uric_acid |
| | Lymphocyte_percentage | Liver function (N=8) | Alanine_transaminase |
| | Mean_platelet_volume | | Albumin |
| | Mean_red_blood_cell_hemoglobin_concentration | | Aspartate_aminotransferase |
| | Mean_red_blood_cell_hemoglobin_content | | Globulin |
| | Mean_red_blood_cell_volume | | Total_bile_acid |
| | Monocyte_absolute_value | | Total_bilirubin |
| | Monocyte_percentage | | Total_protein |
| | Neutrophil_percentage | | White_ball_ratio |
| | Nucleated_red_blood_cell_absolute_value | Thyroid function (N=3) | Free_thyroxine |
| | Nucleated_red_blood_cells | | Nntithyroid_peroxidase_antibody |
| | Platelet_count | | Thyroid_stimulating_hormone |
| | Platelet_hematocrit | Infection (N=5) | Cytomegalovirus_IgM_antibody_quantitative |
| | Platelet_volume_distribution_width | | Herpes_simplex_virus_typeI_IgM_antibody_quantitative |

| | |
|---|---|
| Red_blood_cell_count | Herpes_simplex_virus_typeII_IgM_antibody_quantitative |
| Red_blood_cell_volume_distribution_width_variation | Rubella_virus_IgM_antibody_quantitative |
| Red_blood_cell_volume_distribution_width<br>White_blood_cell | Toxoplasma_IgM_antibody_quantitative |

**ESM Table 3 Clinical characteristics of the study participants**

| | GDM | Nondiabetic control |
|---|---|---|
| **N** | 3,317 | 19,565 |
| **Age (years)** | 31.50±4.23 | 29.32±3.98 |
| **Weight(kg)** | 57.02±8.73 | 54.18±7.59 |
| **BMI (kg/m$^2$)** | 22.74±3.21 | 21.45±2.76 |
| **SBP (mmHg)** | 116.73±13.67 | 114.66±12.95 |
| **DBP (mmHg)** | 68.71±10.30 | 66.84±9.95 |
| **FPG (mmol/L)** | 4.61±0.51 | 4.34±0.32 |
| **OGTT0H (mmol/L)** | 4.80±0.51 | 4.36±0.31 |
| **OGTT1H (mmol/L)** | 9.84±1.39 | 7.28±1.31 |
| **OGTT2H (mmol/L)** | 8.59±1.33 | 6.36±1.00 |

Shown is the mean and the standard deviation of the measurements. N refers to sample size. BMI: Body mass index; SBP: Systolic blood pressure; DBP: Diastolic blood pressure; FPG: Fasting plasma glucose; OGTT0H, 1H and 2H: Oral glucose tolerance test 0 hour, 1 hour and 2 hours.

**ESM Table 4 Statistical test results for clinical characteristics.**

| | GDM | | FPG | | OGTT0H | | 0GTT1H | | OGTT2H | |
|---|---|---|---|---|---|---|---|---|---|---|
| | **OR** | ***P*** | **beta** | ***P*** | **beta** | ***P*** | **beta** | ***P*** | **beta** | ***P*** |
| Age (years) | 1.13 | <0.0001 | 0.03 | <0.0001 | 0.04 | <0.0001 | 0.06 | <0.0001 | 0.06 | <0.0001 |
| Weight(kg) | 1.42 | <0.0001 | 0.20 | <0.0001 | 0.23 | <0.0001 | 0.15 | <0.0001 | 0.13 | <0.0001 |
| BMI (kg/m$^2$) | 1.56 | <0.0001 | 0.20 | <0.001 | 0.24 | <0.0001 | 0.20 | <0.0001 | 0.19 | <0.0001 |
| SBP (mmHg) | 1.01 | <0.0001 | 0.002 | <0.0001 | 0.005 | <0.0001 | 0.005 | <0.0001 | 0.004 | <0.0001 |
| DBP (mmHg) | 1.01 | <0.0001 | 0.002 | <0.0001 | 0.005 | <0.0001 | 0.005 | <0.0001 | 0.004 | <0.0001 |

The qualitative and quantitative variables were analyzed by logictic regression and linear regression analysis without covariate, respectively.GDM: Gestational diabetes mellitus; PGDM: Pre-gestational diabetes mellitus; BMI: Body mass index; SBP: Systolic blood pressure; DBP: Diastolic blood pressure; FPG: Fasting plasma glucose; OGTT0H, 1H and 2H: Oral glucose tolerance test 0 hour, 1 hour and 2 hours.

**ESM Table 5 Heritability of GDM and related quantitative trait and the genetic correlation with T2D.**

| | GDM | FPG | OGTT0H | OGTT1H | OGTT2H |
|---|---|---|---|---|---|
| Sample size | 3317:19565 | 26751 | 24929 | 24931 | 24931 |
| Total Observe scale $h_g^2$ | 0.032(0.019) | 0.053(0.019) | 0.096(0.027) | 0.102(0.022) | 0.078(0.020) |
| Lambda GC | 1.032 | 1.023 | 1.038 | 1.068 | 1.056 |
| Mean Chi^2 | 1.035 | 1.036 | 1.074 | 1.086 | 1.069 |
| Intercept | 1.020(0.006) | 1.007(0.007) | 1.024(0.008) | 1.032(0.008) | 1.027(0.008) |
| Ratio | 0.576(0.176) | 0.178(0.179) | 0.324(0.103) | 0.368(0.091) | 0.390(0.118) |
| Genetic correlation$^{AF}$ | 0.842(0.305) | 0.456(0.188) | 0.523(0.112) | 0.737(0.105) | 0.730(0.118) |
| $P^{AF}$ | 0.006 | 0.015 | 2.77E-06 | 2.71E-12 | 6.02E-10 |
| Genetic correlation$^{A}$ | 0.806(0.240) | 0.307(0.136) | 0.196(0.065) | 0.619(0.079) | 0.695(0.100) |
| $P^{A}$ | 8.0E-04 | 0.024 | 0.003 | 3.94E-15 | 3.88E-12 |
| Genetic correlation$^{E}$ | 0.509(0.205) | 0.145(0.078) | 0.458(0.093) | 0.307(0.064) | 0.352(0.073) |
| $P^{E}$ | 0.013 | 0.062 | 7.75E-07 | 1.37E-06 | 1.28E-06 |

GDM: Gestational diabetes mellitus; FPG: Fasting plasma glucose; OGTT0H, 1H and 2H: Oral glucose tolerance test 0 hour, 1 hour and 2 hours.

[A]:East Asian T2D; [AF]:Asian femaleT2D; [E]:European T2D; Total Observe scale Asian T2D $h_g^2$: 0.078(0.0074); Total Observe scale Asian female T2D $h_g^2$: 0.0664(0.0067);Total Observe scale European T2D $h_g^2$: 0.0692(0.0097). East Asian T2D: the data from the East Asian meta-analyses of 23 T2D GWAS in 433,540 individuals from China, Japan, Korea, Philippines and Malaysia participating in the Asian Genetic Epidemiology Network (AGEN) and Diabetes Meta-analysis of Trans-ethnic Association Studies (DIAMANTE) [17]; European T2D: the data from the meta-analyses of GWAS results from 122 studies and 74,124 T2D cases and 824,006 controls of European ancestry from the DIAMANTE Consortium [18]. T2D data were downloaded from https://t2d.hugeamp.org/.

**ESM Table 6 Heritability of FPG and OGTT2H in the MAGIC consortium.**

| Trait | MAGIC1000G_FG_EAS | MAGIC1000G_FG_EUR | MAGIC1000G_2hGlu_EAS | MAGIC1000G_2hGlu_EUR |
|---|---|---|---|---|
| **Trait sample size** | 35619 | 200622 | 8509 | 63396 |
| **Total_Observed_scale_h2_trait** | 0.020(0.014) | 0.047(0.008) | 0.132(0.053) | 0.058(0.011) |
| **Lambda_GC_trait** | 1.014 | 1.127 | 0.993 | 1.053 |
| **Mean_Chi^2_trait** | 1.031 | 1.242 | 0.994 | 1.070 |
| **Intercept_trait** | 1.019(0.007) | 1.098(0.024) | 0.969(0.006) | 1.019(0.008) |
| **Ratio_trait** | 0.602(0.217) | 0.405(0.100) | Ratio < 0 | 0.276(0.110) |

**ESM Table 7 Replication of the lead SNPs in the NIPT PLUS, BIGCS cohort and meta-analysis.**

| | | | | | | Discovery study | | | NIPT PLUS | | | BIGCS | | | BIGCS and PLUS meta results | | | Discovery versus BIGCS and PLUS meta results | | | |
|---|---|---|---|---|---|---|---|---|---|---|---|---|---|---|---|---|---|---|---|---|---|
| Trait | SNP | CHR | BP | A1 | A2 | BETA | SE | *P* | BETA | SE | *P* | BETA | SE | *P* | BETA | SE | *P* | $P_{diff}$ | Direction | *P* | $P_{het}$ |
| GDM | rs10830963 | 11 | 92975544 | G | C | 0.449 | 0.04 | 4.42E-29 | 0.403 | 0.079 | **3.82E-07** | 0.376 | 0.097 | **1.07E-04** | 0.392 | 0.061 | **1.74E-10** | **0.295** | -- | **7.374E-38** | **0.434** |
| GDM | rs13266634 | 8 | 117172544 | T | C | -0.255 | 0.042 | 1.83E-09 | -0.138 | 0.083 | 9.69E-02 | -0.153 | 0.093 | 9.84E-02 | -0.145 | 0.062 | **1.93E-02** | **0.135** | -- | **3.328E-10** | **0.142** |
| GDM | rs1597916 | 2 | 206947000 | C | G | 0.503 | 0.09 | 1.98E-08 | 0.376 | 0.153 | **1.42E-02** | 0.035 | 0.199 | 8.59E-01 | 0.249 | 0.121 | **4.03E-02** | **0.097** | ++ | **9.807E-09** | **0.092** |
| GDM | rs7766070 | 6 | 20686342 | A | C | 0.235 | 0.034 | 8.10E-12 | 0.13 | 0.071 | 6.86E-02 | 0.198 | 0.1 | **4.83E-02** | 0.153 | 0.058 | **8.55E-03** | **0.189** | ++ | **5.123E-13** | **0.221** |
| OGTT0H | rs10476553 | 5 | 96383142 | C | G | -0.139 | 0.011 | 8.97E-37 | -0.103 | 0.026 | **5.83E-05** | -0.114 | 0.035 | **1.18E-03** | -0.107 | 0.021 | **2.45E-07** | **0.156** | -- | **2.455E-42** | **0.177** |
| OGTT0H | rs10823318 | 10 | 69220168 | T | A | -0.087 | 0.013 | 6.68E-11 | -0.09 | 0.031 | **3.64E-03** | -0.055 | 0.039 | 1.61E-01 | -0.077 | 0.024 | **1.61E-03** | **0.373** | ++ | **4.385E-13** | **0.724** |

| | | | | | | | | | | | | | | | | | | | | |
|---|---|---|---|---|---|---|---|---|---|---|---|---|---|---|---|---|---|---|---|---|
| OGTT0H | rs10974438 | 9 | 4291928 | C | A | 0.071 | 0.012 | 9.74E-10 | 0.053 | 0.028 | 5.61E-02 | 0.075 | 0.035 | **3.34E-02** | 0.062 | 0.022 | **4.81E-03** | **0.373** | -- | **1.756E-11** | **0.700** |
| OGTT0H | rs1912980 | 2 | 60354720 | T | C | -0.077 | 0.012 | 1.15E-10 | -0.084 | 0.028 | **2.62E-03** | -0.052 | 0.037 | 1.56E-01 | -0.072 | 0.022 | **1.13E-03** | **0.392** | -- | **5.13E-13** | **0.848** |
| OGTT0H | rs58048170 | 6 | 20665499 | C | T | 0.062 | 0.011 | 8.95E-09 | -0.001 | 0.025 | 9.57E-01 | 0.055 | 0.035 | 1.16E-01 | 0.018 | 0.021 | 3.82E-01 | **0.067** | -- | **3.681E-08** | **0.060** |
| OGTT0H | rs59858868 | 9 | 687719 | G | A | 0.086 | 0.012 | 1.63E-12 | 0.084 | 0.031 | **6.57E-03** | 0.028 | 0.039 | 4.73E-01 | 0.063 | 0.024 | **9.93E-03** | **0.273** | -- | **7.648E-14** | **0.394** |
| OGTT0H | rs6048209 | 20 | 22590445 | G | A | -0.137 | 0.015 | 4.20E-21 | -0.145 | 0.035 | **3.14E-05** | -0.128 | 0.046 | **5.25E-03** | -0.139 | 0.028 | **5.50E-07** | **0.398** | ++ | **1.258E-26** | **0.953** |
| OGTT0H | rs730497 | 7 | 44184122 | A | G | 0.173 | 0.017 | 3.27E-24 | 0.19 | 0.034 | **2.95E-08** | 0.074 | 0.044 | 9.46E-02 | 0.146 | 0.027 | **6.14E-08** | **0.28** | ++ | **1.542E-30** | **0.407** |
| OGTT0H | rs780094 | 2 | 27518370 | T | C | -0.071 | 0.012 | 1.83E-09 | -0.069 | 0.026 | **7.18E-03** | NA | NA | NA | NA | NA | NA | **NA** | -- | **4.478E-11** | **0.951** |
| OGTT0H | rs7941837 | 11 | 92966500 | T | A | 0.173 | 0.013 | 5.75E-43 | 0.218 | 0.029 | **7.98E-14** | 0.215 | 0.034 | **1.70E-10** | 0.217 | 0.022 | **6.86E-23** | **0.091** | -- | **1.061E-63** | **0.079** |
| OGTT0H | rs853774 | 2 | 168956886 | A | G | -0.147 | 0.014 | 2.88E-26 | -0.161 | 0.029 | **3.54E-08** | -0.084 | 0.034 | **1.46E-02** | -0.129 | 0.022 | **6.88E-09** | **0.312** | -- | **1.425E-33** | **0.489** |
| OGTT1H | rs1001525 | 11 | 35413115 | T | A | -0.085 | 0.014 | 3.56E-09 | -0.013 | 0.035 | 7.06E-01 | -0.055 | 0.04 | 1.67E-01 | -0.032 | 0.026 | 2.32E-01 | **0.08** | ++ | **8.789E-09** | **0.073** |
| OGTT1H | rs118083802 | 6 | 20716830 | C | G | 0.593 | 0.06 | 3.38E-23 | 0.005 | 0.076 | 9.52E-01 | 0.058 | 0.075 | 4.36E-01 | 0.032 | 0.054 | 5.51E-01 | 0.000 | ++ | **1.54E-12** | 0.000 |
| OGTT1H | rs1260334 | 2 | 27525730 | A | C | 0.071 | 0.012 | 8.44E-10 | 0.037 | 0.026 | 1.57E-01 | 0.111 | 0.034 | **1.20E-03** | 0.064 | 0.021 | **2.08E-03** | **0.38** | ++ | **6.762E-12** | **0.755** |
| OGTT1H | rs742762 | 6 | 39078868 | C | A | -0.08 | 0.012 | 5.61E-11 | -0.035 | 0.029 | 2.33E-01 | -0.023 | 0.037 | 5.31E-01 | -0.030 | 0.023 | 1.86E-01 | **0.06** | ++ | **1.508E-10** | **0.053** |
| OGTT1H | rs7683010 | 4 | 57492982 | T | G | 0.059 | 0.011 | 2.79E-08 | 0.068 | 0.025 | **6.79E-03** | NA | NA | NA | NA | NA | NA | **NA** | ++ | **6.836E-10** | **0.739** |
| OGTT1H | rs7941837 | 11 | 92966500 | T | A | 0.218 | 0.012 | 9.59E-70 | 0.234 | 0.029 | **1.99E-15** | 0.227 | 0.034 | **1.79E-11** | 0.231 | 0.022 | **1.81E-25** | **0.35** | -- | **6.04E-94** | **0.607** |
| OGTT1H | rs9650069 | 8 | 117191781 | T | C | -0.115 | 0.018 | 8.65E-11 | -0.085 | 0.031 | **6.36E-03** | -0.049 | 0.035 | 1.61E-01 | -0.069 | 0.023 | **2.99E-03** | **0.118** | -- | **3.268E-12** | **0.120** |
| OGTT2H | rs10830962 | 11 | 92965261 | G | C | 0.146 | 0.013 | 4.74E-30 | 0.146 | 0.029 | **5.87E-07** | 0.129 | 0.034 | **1.92E-04** | 0.139 | 0.022 | **4.70E-10** | **0.384** | -- | **1.363E-38** | **0.785** |
| OGTT2H | rs11975703 | 7 | 44109506 | A | G | -0.079 | 0.013 | 3.05E-09 | -0.075 | 0.031 | **1.49E-02** | -0.091 | 0.04 | **2.36E-02** | -0.081 | 0.024 | **9.38E-04** | **0.398** | -- | **1.115E-11** | **0.960** |
| OGTT2H | rs4746822 | 10 | 69223185 | T | C | 0.114 | 0.013 | 4.33E-18 | 0.152 | 0.031 | **1.42E-06** | 0.14 | 0.039 | **3.44E-04** | 0.147 | 0.025 | **1.88E-09** | **0.196** | ++ | **1.001E-25** | **0.238** |
| OGTT2H | rs7766070 | 6 | 20686342 | A | C | 0.106 | 0.011 | 4.42E-23 | 0.071 | 0.027 | **8.13E-03** | 0.073 | 0.037 | **4.46E-02** | 0.072 | 0.022 | **8.91E-04** | **0.151** | ++ | **4.098E-25** | **0.158** |
| FPG | rs853774 | 2 | 168956886 | A | G | -0.128 | 0.015 | 1.63E-18 | -0.171 | 0.031 | **2.49E-08** | NA | NA | NA | NA | NA | NA | **0.183** | -- | **4.749E-25** | **0.206** |
| FPG | rs1260334 | 2 | 27525730 | A | C | 0.069 | 0.012 | 2.05E-08 | 0.054 | 0.027 | **4.46E-02** | NA | NA | NA | NA | NA | NA | **0.351** | ++ | **2.899E-09** | **0.609** |
| FPG | rs10476553 | 5 | 96383142 | C | G | -0.108 | 0.012 | 7.58E-21 | -0.064 | 0.027 | **1.83E-02** | NA | NA | NA | NA | NA | NA | **0.132** | -- | **1.288E-21** | **0.132** |
| FPG | rs147973072 | 6 | 116933819 | T | C | -1.807 | 0.323 | 2.30E-08 | 0.199 | 0.149 | 1.82E-01 | NA | NA | NA | NA | NA | NA | 0.000 | +- | 0.2613 | 0.000 |
| FPG | rs57501127 | 7 | 44221147 | C | T | 0.499 | 0.063 | 3.41E-15 | 0.198 | 0.084 | **1.80E-02** | NA | NA | NA | NA | NA | NA | 0.007 | -- | **1.207E-14** | 0.004 |
| FPG | rs59858868 | 9 | 687719 | G | A | 0.08 | 0.013 | 4.54E-10 | 0.087 | 0.033 | **7.22E-03** | NA | NA | NA | NA | NA | NA | **0.391** | -- | **1.13E-11** | **0.832** |
| FPG | rs10830962 | 11 | 92965261 | G | C | 0.133 | 0.013 | 6.72E-23 | 0.091 | 0.031 | **3.24E-03** | NA | NA | NA | NA | NA | NA | **0.183** | -- | **1.658E-24** | **0.205** |
| FPG | rs2305483 | 17 | 39986745 | A | C | -1.301 | 0.236 | 3.82E-08 | -0.043 | 0.194 | 8.23E-01 | NA | NA | NA | NA | NA | NA | 0.000 | -- | **0.0002499** | 0.000 |
| FPG | rs6036152 | 20 | 22575955 | C | A | -0.119 | 0.015 | 1.09E-14 | -0.114 | 0.036 | **1.71E-03** | NA | NA | NA | NA | NA | NA | **0.396** | ++ | **7.053E-17** | **0.910** |

*P*: P value of GWAS or meta analysis, values less than 0.05 are bolded. $P_{diff}$: P value for compared the effect estimates from our primary study (the discovery cohort) with those obtained from the BIGCS and PLUS meta-analysis $P_{het}$: P value for heterogeneity in SNP effects through meta-analysis, values greater than 0.05 are bolded.

**ESM Table 8 The information of Lead SNPs about the five traits investigated in the study.**

| Trait | SNP | CHR | BP(GRCh38) | A1 | A2 | FRQ | INFO | BETA | SE | P | GENE | Marker | REGION | EFFECT |
|---|---|---|---|---|---|---|---|---|---|---|---|---|---|---|
| GDM | rs1597916 | 2 | 206947000 | C | G | 0.062 | 0.412 | 0.503 | 0.090 | 1.98E-08 | CPO | novelty | intron_variant | MODIFIER |
| GDM | rs7766070 | 6 | 20686342 | A | C | 0.368 | 0.834 | 0.235 | 0.034 | 8.10E-12 | *CDKAL1* | known | intron_variant | MODIFIER |
| GDM | rs13266634 | 8 | 117172544 | T | C | 0.436 | 0.538 | -0.255 | 0.042 | 1.83E-09 | SLC30A8 | novelty | missense_variant | MODERATE |
| GDM | rs10830963 | 11 | 92975544 | G | C | 0.445 | 0.575 | 0.449 | 0.040 | 4.42E-29 | MTNR1B | known | intron_variant | MODIFIER |
| FPG | rs853774 | 2 | 168956886 | A | G | 0.454 | 0.452 | -0.128 | 0.015 | 1.63E-18 | ABCB11 | known | intron_variant | MODIFIER |
| FPG | rs1260334 | 2 | 27525730 | A | C | 0.488 | 0.651 | 0.069 | 0.012 | 2.05E-08 | GCKR | known | downstream_gene_variant | MODIFIER |
| FPG | rs10476553 | 5 | 96383142 | C | G | 0.347 | 0.816 | -0.108 | 0.012 | 7.58E-21 | LOC101929710 | known | intron_variant | MODIFIER |
| FPG | rs147973072 | 6 | 116933819 | T | C | 0.009 | 0.025 | -1.807 | 0.323 | 2.30E-08 | RFX6 | known | downstream_gene_variant | MODIFIER |
| FPG | rs57501127 | 7 | 44221147 | C | T | 0.047 | 0.136 | 0.499 | 0.063 | 3.41E-15 | CAMK2B | known | intron_variant | MODIFIER |
| FPG | rs59858868 | 9 | 687719 | G | A | 0.261 | 0.788 | 0.080 | 0.013 | 4.54E-10 | KANK1 | known | intron_variant | MODIFIER |
| FPG | rs10830962 | 11 | 92965261 | G | C | 0.451 | 0.534 | 0.133 | 0.013 | 6.72E-23 | MTNR1B | known | upstream_gene_variant | MODIFIER |
| FPG | rs2305483 | 17 | 39986745 | A | C | 0.019 | 0.025 | -1.301 | 0.236 | 3.82E-08 | PSMD3 | novelty | intron_variant | MODIFIER |
| FPG | rs6036152 | 20 | 22575955 | C | A | 0.154 | 0.782 | -0.119 | 0.015 | 1.09E-14 | LINC00261 | known | intron_variant | MODIFIER |
| OGTT0H | rs853774 | 2 | 168956886 | A | G | 0.454 | 0.452 | -0.147 | 0.014 | 2.88E-26 | ABCB11 | known | intron_variant | MODIFIER |
| OGTT0H | rs1912980 | 2 | 60354720 | T | C | 0.333 | 0.685 | -0.077 | 0.012 | 1.15E-10 | MIR4432HG | known | downstream_gene_variant | MODIFIER |
| OGTT0H | rs780094 | 2 | 27518370 | T | C | 0.525 | 0.636 | -0.071 | 0.012 | 1.83E-09 | GCKR | known | intron_variant | MODIFIER |
| OGTT0H | rs10476553 | 5 | 96383142 | C | G | 0.347 | 0.816 | -0.139 | 0.011 | 8.97E-37 | LOC101929710 | known | intron_variant | MODIFIER |
| OGTT0H | rs58048170 | 6 | 20665499 | C | T | 0.380 | 0.834 | 0.062 | 0.011 | 8.95E-09 | CDKAL1 | known | intron_variant | MODIFIER |
| OGTT0H | rs730497 | 7 | 44184122 | A | G | 0.189 | 0.503 | 0.173 | 0.017 | 3.27E-24 | GCK | known | intron_variant | MODIFIER |
| OGTT0H | rs59858868 | 9 | 687719 | G | A | 0.261 | 0.788 | 0.086 | 0.012 | 1.63E-12 | KANK1 | known | intron_variant | MODIFIER |
| OGTT0H | rs10974438 | 9 | 4291928 | C | A | 0.377 | 0.685 | 0.071 | 0.012 | 9.74E-10 | GLIS3 | known | intron_variant | MODIFIER |
| OGTT0H | rs10823318 | 10 | 69220168 | T | A | 0.269 | 0.615 | -0.087 | 0.013 | 6.68E-11 | HKDC1 | novelty | upstream_gene_variant | MODIFIER |
| OGTT0H | rs7941837 | 11 | 92966500 | T | A | 0.438 | 0.560 | 0.173 | 0.013 | 5.75E-43 | MTNR1B | known | upstream_gene_variant | MODIFIER |

| | | | | | | | | | | | | | | |
|---|---|---|---|---|---|---|---|---|---|---|---|---|---|---|
| OGTT0H | rs6048209 | 20 | 22590445 | G | A | 0.149 | 0.801 | -0.137 | 0.015 | 4.20E-21 | FOXA2 | known | upstream_gene_variant | MODIFIER |
| OGTT1H | rs1260334 | 2 | 27525730 | A | C | 0.488 | 0.651 | 0.071 | 0.012 | 8.44E-10 | GCKR | novelty | downstream_gene_variant | MODIFIER |
| OGTT1H | rs7683010 | 4 | 57492982 | T | G | 0.435 | 0.761 | 0.059 | 0.011 | 2.79E-08 | LOC101928851-LOC105377671 | novelty | intergenic_region | MODIFIER |
| OGTT1H | rs118083802 | 6 | 20716830 | C | G | 0.065 | 0.100 | 0.593 | 0.060 | 3.38E-23 | CDKAL1 | novelty | intron_variant | MODIFIER |
| OGTT1H | rs742762 | 6 | 39078868 | C | A | 0.331 | 0.648 | -0.080 | 0.012 | 5.61E-11 | GLP1R | novelty | intron_variant | MODIFIER |
| OGTT1H | rs9650069 | 8 | 117191781 | T | C | 0.279 | 0.339 | -0.115 | 0.018 | 8.65E-11 | SLC30A8-MED30 | novelty | intergenic_region | MODIFIER |
| OGTT1H | rs7941837 | 11 | 92966500 | T | A | 0.438 | 0.560 | 0.218 | 0.012 | 9.59E-70 | MTNR1B | novelty | upstream_gene_variant | MODIFIER |
| OGTT1H | rs1001525 | 11 | 35413115 | T | A | 0.218 | 0.608 | -0.085 | 0.014 | 3.56E-09 | SLC1A2 | novelty | intron_variant | MODIFIER |
| OGTT2H | rs7766070 | 6 | 20686342 | A | C | 0.368 | 0.834 | 0.106 | 0.011 | 4.42E-23 | CDKAL1 | novelty | intron_variant | MODIFIER |
| OGTT2H | rs11975703 | 7 | 44109506 | A | G | 0.252 | 0.645 | -0.079 | 0.013 | 3.05E-09 | MIR4649 | known | upstream_gene_variant | MODIFIER |
| OGTT2H | rs4746822 | 10 | 69223185 | T | C | 0.274 | 0.615 | 0.114 | 0.013 | 4.33E-18 | HKDC1 | novelty | intron_variant | MODIFIER |
| OGTT2H | rs10830962 | 11 | 92965261 | G | C | 0.451 | 0.534 | 0.146 | 0.013 | 4.74E-30 | MTNR1B | novelty | upstream_gene_variant | MODIFIER |

Lead SNP information for Figure 3. CHR: chromosome; BP(GRCh38):position, the version of position is on human GRCh38; A1: the effect allele; FRQ: the frequency of effect allele; INFO indicates the information score in the imputation process; BETA refers to the effect size in the gwas regression model; The ‘Marker’ column indicates whether the SNP was reported in the previous study, reported as known, has not been reported as novel. The ‘REGION’ indicates the SNP function region on chromosome, ‘EFFECT’ column indicates the biological effect of SNP.

**ESM Table 9 Comparison of the 13 significnat loci for FPG and OGTT2H in the MAGIC consortium East Asian population.**

| Trait | SNP | CHR | BP | Effect_allele | Other_allele | This study population | | | | MAGIC_EAS_population | | | | Meta_results | | | |
|---|---|---|---|---|---|---|---|---|---|---|---|---|---|---|---|---|---|
| | | | | | | EAF | BETA | SE | *P* | EAF | BETA | SE | *P* | BETA | SE | *P* | Direction |
| FPG | rs853774 | 2 | 168956886 | A | G | 0.454 | -0.128 | 0.015 | 1.63E-18 | 0.481 | -0.044 | 0.004 | 2.33E-31 | -0.050 | 0.004 | 6.84E-37 | -- |
| FPG | rs1260334 | 2 | 27525730 | A | C | 0.488 | 0.069 | 0.012 | 2.05E-08 | 0.449 | 0.029 | 0.004 | 2.57E-12 | 0.033 | 0.004 | 2.73E-17 | ++ |
| FPG | rs10476553 | 5 | 96383142 | C | G | 0.347 | -0.108 | 0.012 | 7.58E-21 | 0.346 | -0.013 | 0.005 | 1.14E-02 | -0.026 | 0.004 | 1.18E-09 | -- |
| FPG | rs147973072 | 6 | 116933819 | T | C | 0.009 | -1.807 | 0.323 | 2.30E-08 | 0.021 | 0.020 | 0.027 | 3.78E-01 | 0.007 | 0.027 | 7.87E-01 | -+ |
| FPG | rs57501127 | 7 | 44221147 | C | T | 0.047 | 0.499 | 0.063 | 3.41E-15 | NA | NA | NA | NA | 0.499 | 0.063 | 3.23E-15 | -? |
| FPG | rs59858868 | 9 | 687719 | G | A | 0.261 | 0.080 | 0.013 | 4.54E-10 | 0.210 | 0.021 | 0.005 | 8.58E-05 | 0.030 | 0.005 | 1.50E-09 | -- |
| FPG | rs10830962 | 11 | 92965261 | G | C | 0.451 | 0.133 | 0.013 | 6.72E-23 | 0.449 | 0.038 | 0.004 | 1.37E-18 | 0.046 | 0.004 | 6.30E-32 | -- |
| FPG | rs2305483 | 17 | 39986745 | A | C | 0.019 | -1.301 | 0.236 | 3.82E-08 | 0.035 | 0.041 | 0.038 | 2.16E-01 | 0.008 | 0.037 | 8.32E-01 | -+ |
| FPG | rs6036152 | 20 | 22575955 | C | A | 0.154 | -0.119 | 0.015 | 1.09E-14 | 0.154 | -0.045 | 0.006 | 7.67E-14 | -0.054 | 0.005 | 4.23E-24 | ++ |
| OGTT2H | rs7766070 | 6 | 20686342 | A | C | 0.368 | 0.106 | 0.011 | 4.42E-23 | 0.406 | 0.118 | 0.032 | 7.48E-04 | 0.108 | 0.010 | 4.37E-26 | ++ |
| OGTT2H | rs11975703 | 7 | 44109506 | A | G | 0.252 | -0.079 | 0.013 | 3.05E-09 | 0.268 | -0.028 | 0.045 | 4.07E-01 | -0.075 | 0.013 | 4.53E-09 | -- |
| OGTT2H | rs4746822 | 10 | 69223185 | T | C | 0.274 | 0.114 | 0.013 | 4.33E-18 | 0.226 | 0.032 | 0.040 | 2.01E-01 | 0.106 | 0.013 | 2.03E-17 | ++ |
| OGTT2H | rs10830962 | 11 | 92965261 | G | C | 0.451 | 0.146 | 0.013 | 4.74E-30 | 0.449 | -0.002 | 0.031 | 9.19E-01 | 0.125 | 0.012 | 5.52E-26 | -+ |

CHR: chromosome; BP(GRCh38)):position, the version of position is human GRCh38; EAF: effect allele frequency.

### ESM Table 10 Detection specific SNP genetic effects of FPG lead SNPs with OGTT1H and OGTT2H

| | | | | | | | | FPG | | | OGTT1H | | | | OGTT2H | | | |
|---|---|---|---|---|---|---|---|---|---|---|---|---|---|---|---|---|---|---|
| CHR | BP | SNP | A1 | A2 | FRQ | INFO | GENE | BETA | SE | P | BETA | SE | P | Effect_detection $\_P^{FPG\ and\ OGTT1H}$ | BETA | SE | P | Effect_detection $\_P^{FPG\ and\ OGTT2H}$ |
| 2 | 168956886 | rs853774 | A | G | 0.45 | 0.45 | ABCB11 | -0.128 | 0.015 | 1.63E-18 | -0.003 | 0.014 | 8.11E-01 | 1.30E-09 | -0.027 | 0.014 | 5.06E-02 | 1.31E-06 |
| 2 | 27525730 | rs1260334 | A | C | 0.49 | 0.65 | GCKR | 0.069 | 0.012 | 2.05E-08 | 0.071 | 0.012 | 8.44E-10 | 3.97E-01 | 0.019 | 0.012 | 1.00E-01 | 5.38E-03 |
| 5 | 96383142 | rs10476553 | C | G | 0.35 | 0.82 | LOC101929710 | -0.108 | 0.012 | 7.58E-21 | -0.046 | 0.011 | 2.05E-05 | 1.76E-04 | -0.038 | 0.011 | 4.60E-04 | 2.76E-05 |
| 6 | 116933819 | rs147973072 | T | C | RFX6 | 0.009 | 0.025 | -1.807 | 0.323 | 2.30E-08 | -1.609 | 0.307 | 1.54E-07 | 3.61E-01 | 0.095 | 0.307 | 7.57E-01 | 4.38E-05 |
| 7 | 44221147 | rs57501127 | C | T | CAMK2B | 0.047 | 0.136 | 0.499 | 0.063 | 3.41E-15 | 0.194 | 0.059 | 1.04E-03 | 8.09E-04 | 0.222 | 0.060 | 2.27E-04 | 2.52E-03 |
| 9 | 687719 | rs59858868 | G | A | 0.26 | 0.79 | KANK1 | 0.080 | 0.013 | 4.54E-10 | -0.023 | 0.012 | 5.10E-02 | 1.14E-08 | -0.023 | 0.012 | 5.63E-02 | 1.53E-08 |
| 11 | 92965261 | rs10830962 | G | C | 0.45 | 0.53 | MTNR1B | 0.133 | 0.013 | 6.72E-23 | 0.222 | 0.013 | 1.12E-69 | 3.52E-06 | 0.146 | 0.013 | 4.74E-30 | 3.16E-01 |
| 17 | 39986745 | rs2305483 | A | C | PSMD3 | 0.019 | 0.025 | -1.301 | 0.236 | 3.82E-08 | 0.241 | 0.220 | 2.73E-01 | 4.41E-06 | -0.110 | 0.224 | 6.21E-01 | 4.91E-04 |
| 20 | 22575955 | rs6036152 | C | A | 0.15 | 0.78 | LINC00261 | -0.119 | 0.015 | 1.09E-14 | -0.050 | 0.014 | 5.23E-04 | 1.81E-03 | -0.064 | 0.015 | 1.22E-05 | 1.36E-02 |

Effect_detection $\_P^{FPG\ and\ OGTT1H}$: two-sample *t*-test *P*-value of difference in specific SNP genetic effects between FPG and OGTT1H GWAS; Effect_detection $\_P^{FPG\ and\ OGTT2H}$: two-sample *t*-test *P*-value of difference in specific SNP genetic effects between FPG and OGTT2H. A *P*-value less than 0.05 suggests a statistically significant difference in the effect of the Single Nucleotide Polymorphism (SNP) . Details of the two-sample *t*-test can be referred to Supplementary Notes.

### ESM Table 11 MR results of GDM and 4 quantitative glycemic traits and biomarkers.

| exposure | outcome | method | nsnp | b | se | pval | method_het | Q | Q_pval | egger_intercept | pleio_se | pleio_p |
|---|---|---|---|---|---|---|---|---|---|---|---|---|
| Absolute_neutrophils | GDM | MR Egger | 17 | -0.283 | 0.079 | 2.84E-03 | MR Egger | 10.864 | 0.762 | 0.015 | 0.031 | 0.629 |
| Absolute_neutrophils | GDM | Weighted median | 17 | -0.276 | 0.081 | 6.75E-04 | MR Egger | 10.864 | 0.762 | 0.015 | 0.031 | 0.629 |
| Absolute_neutrophils | GDM | Inverse variance weighted | 17 | -0.260 | 0.065 | 6.57E-05 | MR Egger | 10.864 | 0.762 | 0.015 | 0.031 | 0.629 |
| Absolute_neutrophils | GDM | Simple mode | 17 | -0.271 | 0.111 | 2.65E-02 | MR Egger | 10.864 | 0.762 | 0.015 | 0.031 | 0.629 |
| Absolute_neutrophils | GDM | Weighted mode | 17 | -0.292 | 0.086 | 3.71E-03 | MR Egger | 10.864 | 0.762 | 0.015 | 0.031 | 0.629 |

| | | | | | | | | | | | | |
|---|---|---|---|---|---|---|---|---|---|---|---|---|
| Lymphocyte_percentage | GDM | MR Egger | 9 | 0.512 | 0.129 | 5.30E-03 | Inverse variance weighted | 3.300 | 0.914 | -0.031 | 0.029 | 0.332 |
| Lymphocyte_percentage | GDM | Weighted median | 9 | 0.471 | 0.127 | 2.01E-04 | Inverse variance weighted | 3.300 | 0.914 | -0.031 | 0.029 | 0.332 |
| Lymphocyte_percentage | GDM | Inverse variance weighted | 9 | 0.436 | 0.106 | 3.74E-05 | Inverse variance weighted | 3.300 | 0.914 | -0.031 | 0.029 | 0.332 |
| Lymphocyte_percentage | GDM | Simple mode | 9 | 0.491 | 0.161 | 1.57E-02 | Inverse variance weighted | 3.300 | 0.914 | -0.031 | 0.029 | 0.332 |
| Lymphocyte_percentage | GDM | Weighted mode | 9 | 0.491 | 0.158 | 1.44E-02 | Inverse variance weighted | 3.300 | 0.914 | -0.031 | 0.029 | 0.332 |
| Neutrophil_percentage | GDM | MR Egger | 10 | -0.481 | 0.121 | 4.14E-03 | MR Egger | 8.211 | 0.413 | 0.020 | 0.032 | 0.550 |
| Neutrophil_percentage | GDM | Weighted median | 10 | -0.447 | 0.126 | 3.82E-04 | MR Egger | 8.211 | 0.413 | 0.020 | 0.032 | 0.550 |
| Neutrophil_percentage | GDM | Inverse variance weighted | 10 | -0.438 | 0.099 | 9.47E-06 | MR Egger | 8.211 | 0.413 | 0.020 | 0.032 | 0.550 |
| Neutrophil_percentage | GDM | Simple mode | 10 | -0.470 | 0.155 | 1.42E-02 | MR Egger | 8.211 | 0.413 | 0.020 | 0.032 | 0.550 |
| Neutrophil_percentage | GDM | Weighted mode | 10 | -0.470 | 0.148 | 1.14E-02 | MR Egger | 8.211 | 0.413 | 0.020 | 0.032 | 0.550 |
| White_blood_cell | GDM | MR Egger | 23 | -0.249 | 0.071 | 1.99E-03 | MR Egger | 13.238 | 0.900 | 0.009 | 0.019 | 0.645 |
| White_blood_cell | GDM | Weighted median | 23 | -0.268 | 0.079 | 6.51E-04 | MR Egger | 13.238 | 0.900 | 0.009 | 0.019 | 0.645 |
| White_blood_cell | GDM | Inverse variance weighted | 23 | -0.233 | 0.061 | 1.38E-04 | MR Egger | 13.238 | 0.900 | 0.009 | 0.019 | 0.645 |
| White_blood_cell | GDM | Simple mode | 23 | -0.257 | 0.111 | 2.98E-02 | MR Egger | 13.238 | 0.900 | 0.009 | 0.019 | 0.645 |
| White_blood_cell | GDM | Weighted mode | 23 | -0.281 | 0.080 | 2.04E-03 | MR Egger | 13.238 | 0.900 | 0.009 | 0.019 | 0.645 |

Method_het and Q_pval refer to the model of heterogeneity test and p value of statistic test; egger_intercept, pleio_se,pleio_p refer to the intercept,se and P value of pleiotropy test.

**ESM Table 12 Instrumental variables of biomarkers as exposure and GDM as outcome.**

| exposure | outcome | SNP | effect_allele | other_allele | eaf | beta.exposure | se.exposure | pval.exposure | samplesize.exposure | R2 | F | beta.outcome | se.outcome | pval.outcome | samplesize.outcome |
|---|---|---|---|---|---|---|---|---|---|---|---|---|---|---|---|
| Absolute_neutrophils | GDM | rs116877864 | C | G | 0.02 | -2.75 | 0.15 | 8.29E-78 | 28541 | 0.012 | 350.80 | 1.07 | 0.54 | 0.05 | 22882 |
| Absolute_neutrophils | GDM | rs117264195 | C | T | 0.04 | -0.71 | 0.10 | 1.11E-12 | 28541 | 0.002 | 50.68 | -0.19 | 0.37 | 0.61 | 22882 |
| Absolute_neutrophils | GDM | rs118092418 | A | G | 0.04 | 1.29 | 0.08 | 2.20E-52 | 28541 | 0.008 | 232.93 | -0.11 | 0.31 | 0.73 | 22882 |
| Absolute_neutrophils | GDM | rs141690080 | G | A | 0.02 | 2.07 | 0.23 | 6.47E-19 | 28541 | 0.003 | 79.03 | 0.66 | 0.85 | 0.44 | 22882 |
| Absolute_neutrophils | GDM | rs142327763 | G | C | 0.04 | 1.58 | 0.09 | 2.04E-74 | 28541 | 0.012 | 335.05 | -0.43 | 0.32 | 0.18 | 22882 |
| Absolute_neutrophils | GDM | rs142772971 | G | T | 0.02 | 0.78 | 0.11 | 1.94E-13 | 28541 | 0.002 | 54.12 | -0.12 | 0.39 | 0.77 | 22882 |
| Absolute_neutrophils | GDM | rs143437186 | T | C | 0.01 | 1.85 | 0.33 | 2.83E-08 | 28541 | 0.001 | 30.84 | -1.84 | 1.23 | 0.13 | 22882 |

| | | | | | | | | | | | | | | | |
|---|---|---|---|---|---|---|---|---|---|---|---|---|---|---|---|
| Absolute_neutrophils | GDM | rs144745936 | A | T | 0.02 | 3.17 | 0.16 | 1.08E-89 | 28541 | 0.014 | 406.12 | -1.03 | 0.59 | 0.08 | 22882 |
| Absolute_neutrophils | GDM | rs181359740 | T | C | 0.03 | 2.88 | 0.12 | 1.68E-117 | 28541 | 0.018 | 536.02 | -0.76 | 0.46 | 0.10 | 22882 |
| Absolute_neutrophils | GDM | rs184338334 | G | A | 0.02 | 0.81 | 0.14 | 2.28E-08 | 28541 | 0.001 | 31.25 | -0.96 | 0.53 | 0.07 | 22882 |
| Absolute_neutrophils | GDM | rs191558349 | A | G | 0.04 | -0.70 | 0.11 | 1.98E-10 | 28541 | 0.001 | 40.51 | -0.37 | 0.40 | 0.36 | 22882 |
| Absolute_neutrophils | GDM | rs2305483 | A | C | 0.02 | 3.73 | 0.20 | 5.46E-80 | 28541 | 0.012 | 360.94 | -1.20 | 0.73 | 0.10 | 22882 |
| Absolute_neutrophils | GDM | rs7021 | A | T | 0.46 | 0.21 | 0.01 | 6.29E-120 | 28541 | 0.019 | 547.39 | -0.06 | 0.03 | 0.08 | 22882 |
| Absolute_neutrophils | GDM | rs76714842 | A | G | 0.19 | 0.09 | 0.01 | 3.72E-14 | 28541 | 0.002 | 57.37 | 0.00 | 0.04 | 0.93 | 22882 |
| Absolute_neutrophils | GDM | rs77115032 | G | A | 0.02 | 0.72 | 0.10 | 3.59E-12 | 28541 | 0.002 | 48.38 | -0.22 | 0.38 | 0.55 | 22882 |
| Absolute_neutrophils | GDM | rs77474927 | T | C | 0.03 | 0.62 | 0.12 | 8.69E-08 | 28541 | 0.001 | 28.66 | -0.55 | 0.43 | 0.19 | 22882 |
| Absolute_neutrophils | GDM | rs78431470 | T | G | 0.09 | -0.19 | 0.04 | 1.02E-07 | 28541 | 0.001 | 28.35 | -0.06 | 0.14 | 0.64 | 22882 |
| Lymphocyte_percentage | GDM | rs116877864 | C | G | 0.02 | 1.78 | 0.15 | 4.66E-33 | 28341 | 0.005 | 143.82 | 1.07 | 0.54 | 0.05 | 22882 |
| Lymphocyte_percentage | GDM | rs118092418 | A | G | 0.04 | -0.82 | 0.08 | 6.32E-22 | 28341 | 0.003 | 92.77 | -0.11 | 0.31 | 0.73 | 22882 |
| Lymphocyte_percentage | GDM | rs142327763 | G | C | 0.04 | -0.94 | 0.09 | 4.49E-27 | 28341 | 0.004 | 116.35 | -0.43 | 0.32 | 0.18 | 22882 |
| Lymphocyte_percentage | GDM | rs150194598 | A | G | 0.03 | -2.66 | 0.18 | 5.41E-48 | 28341 | 0.007 | 212.65 | -1.47 | 0.67 | 0.03 | 22882 |
| Lymphocyte_percentage | GDM | rs16851441 | G | A | 0.24 | 0.07 | 0.01 | 1.47E-08 | 28341 | 0.001 | 32.11 | -0.03 | 0.04 | 0.48 | 22882 |
| Lymphocyte_percentage | GDM | rs181359740 | T | C | 0.03 | -1.79 | 0.13 | 7.58E-46 | 28341 | 0.007 | 202.73 | -0.76 | 0.46 | 0.10 | 22882 |
| Lymphocyte_percentage | GDM | rs2305483 | A | C | 0.02 | -2.42 | 0.20 | 5.19E-34 | 28341 | 0.005 | 148.20 | -1.20 | 0.73 | 0.10 | 22882 |
| Lymphocyte_percentage | GDM | rs4795420 | G | A | 0.46 | -0.14 | 0.01 | 3.56E-48 | 28341 | 0.007 | 213.49 | -0.06 | 0.03 | 0.06 | 22882 |
| Lymphocyte_percentage | GDM | rs7752721 | T | C | 0.07 | 0.09 | 0.02 | 2.72E-08 | 28341 | 0.001 | 30.91 | 0.02 | 0.06 | 0.79 | 22882 |
| Neutrophil_percentage | GDM | rs116877864 | C | G | 0.02 | -1.86 | 0.15 | 1.10E-35 | 28456 | 0.005 | 155.90 | 1.07 | 0.54 | 0.05 | 22882 |
| Neutrophil_percentage | GDM | rs118092418 | A | G | 0.04 | 0.86 | 0.08 | 2.13E-24 | 28456 | 0.004 | 104.08 | -0.11 | 0.31 | 0.73 | 22882 |
| Neutrophil_percentage | GDM | rs141690080 | G | A | 0.02 | 1.34 | 0.23 | 1.02E-08 | 28456 | 0.001 | 32.83 | 0.66 | 0.85 | 0.44 | 22882 |
| Neutrophil_percentage | GDM | rs142327763 | G | C | 0.04 | 1.00 | 0.09 | 1.12E-30 | 28456 | 0.005 | 132.88 | -0.43 | 0.32 | 0.18 | 22882 |
| Neutrophil_percentage | GDM | rs142470396 | G | A | 0.04 | -0.39 | 0.06 | 9.33E-10 | 28456 | 0.001 | 37.48 | 0.62 | 0.22 | 0.01 | 22882 |
| Neutrophil_percentage | GDM | rs150194598 | A | G | 0.03 | 2.82 | 0.18 | 5.38E-54 | 28456 | 0.008 | 240.38 | -1.47 | 0.67 | 0.03 | 22882 |
| Neutrophil_percentage | GDM | rs181359740 | T | C | 0.03 | 1.91 | 0.13 | 4.20E-52 | 28456 | 0.008 | 231.63 | -0.76 | 0.46 | 0.10 | 22882 |
| Neutrophil_percentage | GDM | rs2305483 | A | C | 0.02 | 2.49 | 0.20 | 4.11E-36 | 28456 | 0.006 | 157.87 | -1.20 | 0.73 | 0.10 | 22882 |
| Neutrophil_percentage | GDM | rs4795420 | G | A | 0.46 | 0.14 | 0.01 | 3.51E-54 | 28456 | 0.008 | 241.24 | -0.06 | 0.03 | 0.06 | 22882 |
| Neutrophil_percentage | GDM | rs80347875 | T | A | 0.20 | -0.07 | 0.01 | 1.26E-08 | 28456 | 0.001 | 32.42 | -0.02 | 0.04 | 0.65 | 22882 |

| | | | | | | | | | | | | | | | |
|---|---|---|---|---|---|---|---|---|---|---|---|---|---|---|---|
| White_blood_cell | GDM | rs116877864 | C | G | 0.02 | -2.87 | 0.15 | 3.58E-85 | 28477 | 0.013 | 385.07 | 1.07 | 0.54 | 0.05 | 22882 |
| White_blood_cell | GDM | rs117264195 | C | T | 0.04 | -0.78 | 0.10 | 3.72E-15 | 28477 | 0.002 | 61.91 | -0.19 | 0.37 | 0.61 | 22882 |
| White_blood_cell | GDM | rs118092418 | A | G | 0.04 | 1.32 | 0.08 | 1.43E-55 | 28477 | 0.009 | 247.67 | -0.11 | 0.31 | 0.73 | 22882 |
| White_blood_cell | GDM | rs12672925 | T | C | 0.06 | -0.22 | 0.04 | 1.27E-07 | 28477 | 0.001 | 27.93 | -0.04 | 0.15 | 0.81 | 22882 |
| White_blood_cell | GDM | rs141690080 | G | A | 0.02 | 2.02 | 0.23 | 2.67E-18 | 28477 | 0.003 | 76.22 | 0.66 | 0.85 | 0.44 | 22882 |
| White_blood_cell | GDM | rs142327763 | G | C | 0.04 | 1.61 | 0.09 | 8.17E-78 | 28477 | 0.012 | 350.84 | -0.43 | 0.32 | 0.18 | 22882 |
| White_blood_cell | GDM | rs142772971 | G | T | 0.02 | 0.82 | 0.11 | 8.27E-15 | 28477 | 0.002 | 60.33 | -0.12 | 0.39 | 0.77 | 22882 |
| White_blood_cell | GDM | rs143437186 | T | C | 0.01 | 1.91 | 0.33 | 9.55E-09 | 28477 | 0.001 | 32.95 | -1.84 | 1.23 | 0.13 | 22882 |
| White_blood_cell | GDM | rs144745936 | A | T | 0.02 | 3.27 | 0.16 | 7.09E-96 | 28477 | 0.015 | 434.95 | -1.03 | 0.59 | 0.08 | 22882 |
| White_blood_cell | GDM | rs181359740 | T | C | 0.03 | 2.94 | 0.12 | 4.28E-124 | 28477 | 0.020 | 566.94 | -0.76 | 0.46 | 0.10 | 22882 |
| White_blood_cell | GDM | rs184338334 | G | A | 0.02 | 0.83 | 0.14 | 8.25E-09 | 28477 | 0.001 | 33.23 | -0.96 | 0.53 | 0.07 | 22882 |
| White_blood_cell | GDM | rs191558349 | A | G | 0.04 | -0.77 | 0.11 | 1.40E-12 | 28477 | 0.002 | 50.23 | -0.37 | 0.40 | 0.36 | 22882 |
| White_blood_cell | GDM | rs2299002 | C | A | 0.27 | -0.06 | 0.01 | 1.67E-09 | 28477 | 0.001 | 36.34 | 0.00 | 0.04 | 0.98 | 22882 |
| White_blood_cell | GDM | rs2305483 | A | C | 0.02 | 3.95 | 0.20 | 1.69E-90 | 28477 | 0.014 | 409.88 | -1.20 | 0.73 | 0.10 | 22882 |
| White_blood_cell | GDM | rs3760531 | C | G | 0.02 | -0.89 | 0.15 | 1.68E-09 | 28477 | 0.001 | 36.34 | -0.03 | 0.55 | 0.96 | 22882 |
| White_blood_cell | GDM | rs6989110 | C | T | 0.30 | -0.07 | 0.01 | 1.11E-08 | 28477 | 0.001 | 32.66 | 0.02 | 0.04 | 0.59 | 22882 |
| White_blood_cell | GDM | rs7021 | A | T | 0.46 | 0.22 | 0.01 | 1.79E-129 | 28477 | 0.020 | 592.17 | -0.06 | 0.03 | 0.08 | 22882 |
| White_blood_cell | GDM | rs76677887 | C | T | 0.20 | 0.09 | 0.01 | 9.69E-16 | 28477 | 0.002 | 64.56 | 0.00 | 0.04 | 0.99 | 22882 |
| White_blood_cell | GDM | rs7692 | A | C | 0.31 | -0.06 | 0.01 | 1.50E-08 | 28477 | 0.001 | 32.07 | 0.01 | 0.04 | 0.76 | 22882 |
| White_blood_cell | GDM | rs77115032 | G | A | 0.02 | 0.74 | 0.10 | 5.20E-13 | 28477 | 0.002 | 52.17 | -0.22 | 0.38 | 0.55 | 22882 |
| White_blood_cell | GDM | rs77474927 | T | C | 0.03 | 0.64 | 0.12 | 3.76E-08 | 28477 | 0.001 | 30.28 | -0.55 | 0.43 | 0.19 | 22882 |
| White_blood_cell | GDM | rs78431470 | T | G | 0.09 | -0.21 | 0.04 | 7.81E-09 | 28477 | 0.001 | 33.34 | -0.06 | 0.14 | 0.64 | 22882 |
| White_blood_cell | GDM | rs79540179 | C | T | 0.01 | -3.30 | 0.43 | 3.24E-14 | 28477 | 0.002 | 57.64 | -1.13 | 1.60 | 0.48 | 22882 |

$$F = \frac{N - k - 1}{k} \times \frac{R^2}{1 - R^2}$$

$$R^2 = \frac{2 * \beta^2 * MAF * (1 - MAF)}{2 * \beta^2 * MAF * (1 - MAF) + se^2 * 2 * N * MAF * (1 - MAF)}$$

**ESM Table 13 The power of Mendelian Randomization(MR) analyses.**

| Exposure | Outcome | Sample size | α | K | OR | $R^2$ | Power | F-statistics |
|---|---|---|---|---|---|---|---|---|
| Absolute_neutrophils | GDM | 22882 | 0.05 | 0.14 | 0.77 | 0.116 | 0.99 | 3003.62 |
| Lymphocyte_percentage | GDM | 22882 | 0.05 | 0.14 | 1.55 | 0.042 | 1 | 1004.18 |
| Neutrophil_percentage | GDM | 22882 | 0.05 | 0.14 | 0.65 | 0.048 | 0.99 | 1154.71 |
| White_blood_cell | GDM | 22882 | 0.05 | 0.14 | 0.79 | 0.128 | 0.98 | 3359.83 |

α: type I error rate
K: proportion of cases in the study
OR: true odds ratio of the outcome variable per standard deviation of the exposure variable
$R^2$: Proportion of variance explained for the association between the SNP or allele score (Z) and the exposure variable (X)
F-statistic: the strength of the instrument

**ESM Table 14 MR results of GDM and 4 quantitative glycemic traits and 4 biomarkers use meta data with PLUS cohort.**

| exposure | outcome | method | nsnp | b | se | pval | method_het | Q | Q_df | Q_pval | egger_intercept | pleio_se | pleio_p |
|---|---|---|---|---|---|---|---|---|---|---|---|---|---|
| Absolute_neutrophils | GDM | MR Egger | 16 | -0.243 | 0.082 | 1.05E-02 | MR Egger | 11.00705288 | 14 | 0.685 | 0.002 | 0.014 | 0.866 |
| Absolute_neutrophils | GDM | Weighted median | 16 | -0.219 | 0.086 | 1.10E-02 | MR Egger | 11.00705288 | 14 | 0.685 | 0.002 | 0.014 | 0.866 |
| Absolute_neutrophils | GDM | Inverse variance weighted | 16 | -0.235 | 0.067 | 4.44E-04 | MR Egger | 11.00705288 | 14 | 0.685 | 0.002 | 0.014 | 0.866 |
| Absolute_neutrophils | GDM | Simple mode | 16 | -0.191 | 0.121 | 1.36E-01 | MR Egger | 11.00705288 | 14 | 0.685 | 0.002 | 0.014 | 0.866 |
| Absolute_neutrophils | GDM | Weighted mode | 16 | -0.215 | 0.087 | 2.65E-02 | MR Egger | 11.00705288 | 14 | 0.685 | 0.002 | 0.014 | 0.866 |
| Lymphocyte_percentage | GDM | MR Egger | 12 | 0.558 | 0.154 | 4.71E-03 | Inverse variance weighted | 17.43531351 | 11 | 0.096 | -0.059 | 0.021 | 0.019 |
| Lymphocyte_percentage | GDM | Weighted median | 12 | 0.352 | 0.144 | 1.48E-02 | Inverse variance weighted | 17.43531351 | 11 | 0.096 | -0.059 | 0.021 | 0.019 |
| Lymphocyte_percentage | GDM | Inverse variance weighted | 12 | 0.247 | 0.135 | 6.69E-02 | Inverse variance weighted | 17.43531351 | 11 | 0.096 | -0.059 | 0.021 | 0.019 |
| Lymphocyte_percentage | GDM | Simple mode | 12 | 0.349 | 0.188 | 9.01E-02 | Inverse variance weighted | 17.43531351 | 11 | 0.096 | -0.059 | 0.021 | 0.019 |
| Lymphocyte_percentage | GDM | Weighted mode | 12 | 0.362 | 0.161 | 4.58E-02 | Inverse variance weighted | 17.43531351 | 11 | 0.096 | -0.059 | 0.021 | 0.019 |
| Neutrophil_percentage | GDM | MR Egger | 13 | -0.555 | 0.147 | 3.05E-03 | Inverse variance weighted | 14.62092434 | 12 | 0.263 | 0.047 | 0.021 | 0.044 |
| Neutrophil_percentage | GDM | Weighted median | 13 | -0.349 | 0.136 | 1.06E-02 | Inverse variance weighted | 14.62092434 | 12 | 0.263 | 0.047 | 0.021 | 0.044 |
| Neutrophil_percentage | GDM | Inverse variance weighted | 13 | -0.315 | 0.112 | 5.06E-03 | Inverse variance weighted | 14.62092434 | 12 | 0.263 | 0.047 | 0.021 | 0.044 |

| | | | | | | | | | | | | | |
|---|---|---|---|---|---|---|---|---|---|---|---|---|---|
| Neutrophil_percentage | GDM | Simple mode | 13 | -0.397 | 0.202 | 7.31E-02 | Inverse variance weighted | 14.62092434 | 12 | 0.263 | 0.047 | 0.021 | 0.044 |
| Neutrophil_percentage | GDM | Weighted mode | 13 | -0.377 | 0.147 | 2.45E-02 | Inverse variance weighted | 14.62092434 | 12 | 0.263 | 0.047 | 0.021 | 0.044 |
| White_blood_cell | GDM | MR Egger | 19 | -0.211 | 0.079 | 1.57E-02 | MR Egger | 13.52139763 | 17 | 0.701 | 0.000 | 0.014 | 0.979 |
| White_blood_cell | GDM | Weighted median | 19 | -0.211 | 0.085 | 1.36E-02 | MR Egger | 13.52139763 | 17 | 0.701 | 0.000 | 0.014 | 0.979 |
| White_blood_cell | GDM | Inverse variance weighted | 19 | -0.210 | 0.063 | 8.97E-04 | MR Egger | 13.52139763 | 17 | 0.701 | 0.000 | 0.014 | 0.979 |
| White_blood_cell | GDM | Simple mode | 19 | -0.161 | 0.131 | 2.34E-01 | MR Egger | 13.52139763 | 17 | 0.701 | 0.000 | 0.014 | 0.979 |
| White_blood_cell | GDM | Weighted mode | 19 | -0.213 | 0.087 | 2.46E-02 | MR Egger | 13.52139763 | 17 | 0.701 | 0.000 | 0.014 | 0.979 |
| Absolute_neutrophils | FPG | MR Egger | 16 | -0.256 | 0.029 | 3.90E-07 | MR Egger | 14.10923329 | 14 | 0.442 | 0.004 | 0.005 | 0.392 |
| Absolute_neutrophils | FPG | Weighted median | 16 | -0.236 | 0.034 | 3.12E-12 | MR Egger | 14.10923329 | 14 | 0.442 | 0.004 | 0.005 | 0.392 |
| Absolute_neutrophils | FPG | Inverse variance weighted | 16 | -0.241 | 0.023 | 1.91E-25 | MR Egger | 14.10923329 | 14 | 0.442 | 0.004 | 0.005 | 0.392 |
| Absolute_neutrophils | FPG | Simple mode | 16 | -0.265 | 0.050 | 8.88E-05 | MR Egger | 14.10923329 | 14 | 0.442 | 0.004 | 0.005 | 0.392 |
| Absolute_neutrophils | FPG | Weighted mode | 16 | -0.255 | 0.038 | 7.04E-06 | MR Egger | 14.10923329 | 14 | 0.442 | 0.004 | 0.005 | 0.392 |
| Lymphocyte_percentage | FPG | MR Egger | 12 | 0.457 | 0.067 | 4.59E-05 | Inverse variance weighted | 27.04584468 | 11 | 0.005 | -0.025 | 0.009 | 0.021 |
| Lymphocyte_percentage | FPG | Weighted median | 12 | 0.372 | 0.055 | 1.41E-11 | Inverse variance weighted | 27.04584468 | 11 | 0.005 | -0.025 | 0.009 | 0.021 |
| Lymphocyte_percentage | FPG | Inverse variance weighted | 12 | 0.324 | 0.058 | 2.22E-08 | Inverse variance weighted | 27.04584468 | 11 | 0.005 | -0.025 | 0.009 | 0.021 |
| Lymphocyte_percentage | FPG | Simple mode | 12 | 0.379 | 0.088 | 1.19E-03 | Inverse variance weighted | 27.04584468 | 11 | 0.005 | -0.025 | 0.009 | 0.021 |
| Lymphocyte_percentage | FPG | Weighted mode | 12 | 0.389 | 0.062 | 6.45E-05 | Inverse variance weighted | 27.04584468 | 11 | 0.005 | -0.025 | 0.009 | 0.021 |
| Neutrophil_percentage | FPG | MR Egger | 13 | -0.422 | 0.056 | 1.18E-05 | Inverse variance weighted | 18.83611261 | 12 | 0.093 | 0.017 | 0.008 | 0.054 |
| Neutrophil_percentage | FPG | Weighted median | 13 | -0.359 | 0.050 | 5.20E-13 | Inverse variance weighted | 18.83611261 | 12 | 0.093 | 0.017 | 0.008 | 0.054 |
| Neutrophil_percentage | FPG | Inverse variance weighted | 13 | -0.333 | 0.044 | 3.30E-14 | Inverse variance weighted | 18.83611261 | 12 | 0.093 | 0.017 | 0.008 | 0.054 |
| Neutrophil_percentage | FPG | Simple mode | 13 | -0.393 | 0.088 | 7.31E-04 | Inverse variance weighted | 18.83611261 | 12 | 0.093 | 0.017 | 0.008 | 0.054 |
| Neutrophil_percentage | FPG | Weighted mode | 13 | -0.389 | 0.062 | 4.33E-05 | Inverse variance weighted | 18.83611261 | 12 | 0.093 | 0.017 | 0.008 | 0.054 |
| White_blood_cell | FPG | MR Egger | 19 | -0.227 | 0.028 | 2.48E-07 | MR Egger | 17.10533285 | 17 | 0.447 | 0.003 | 0.005 | 0.581 |
| White_blood_cell | FPG | Weighted median | 19 | -0.221 | 0.032 | 3.05E-12 | MR Egger | 17.10533285 | 17 | 0.447 | 0.003 | 0.005 | 0.581 |
| White_blood_cell | FPG | Inverse variance weighted | 19 | -0.217 | 0.022 | 2.76E-23 | MR Egger | 17.10533285 | 17 | 0.447 | 0.003 | 0.005 | 0.581 |
| White_blood_cell | FPG | Simple mode | 19 | -0.252 | 0.051 | 1.04E-04 | MR Egger | 17.10533285 | 17 | 0.447 | 0.003 | 0.005 | 0.581 |
| White_blood_cell | FPG | Weighted mode | 19 | -0.231 | 0.034 | 2.49E-06 | MR Egger | 17.10533285 | 17 | 0.447 | 0.003 | 0.005 | 0.581 |
| Absolute_neutrophils | OGTT0H | MR Egger | 16 | -0.197 | 0.031 | 1.62E-05 | Inverse variance weighted | 21.12330653 | 15 | 0.133 | -0.008 | 0.005 | 0.128 |
| Absolute_neutrophils | OGTT0H | Weighted median | 16 | -0.190 | 0.030 | 1.70E-10 | Inverse variance weighted | 21.12330653 | 15 | 0.133 | -0.008 | 0.005 | 0.128 |

| | | | | | | | | | | | | | |
|---|---|---|---|---|---|---|---|---|---|---|---|---|---|
| Absolute_neutrophils | OGTT0H | Inverse variance weighted | 16 | -0.226 | 0.026 | 3.28E-18 | Inverse variance weighted | 21.12330653 | 15 | 0.133 | -0.008 | 0.005 | 0.128 |
| Absolute_neutrophils | OGTT0H | Simple mode | 16 | -0.249 | 0.044 | 5.08E-05 | Inverse variance weighted | 21.12330653 | 15 | 0.133 | -0.008 | 0.005 | 0.128 |
| Absolute_neutrophils | OGTT0H | Weighted mode | 16 | -0.197 | 0.030 | 9.50E-06 | Inverse variance weighted | 21.12330653 | 15 | 0.133 | -0.008 | 0.005 | 0.128 |
| Lymphocyte_percentage | OGTT0H | MR Egger | 12 | 0.413 | 0.065 | 8.40E-05 | Inverse variance weighted | 33.35787269 | 11 | 0.000 | -0.029 | 0.009 | 0.008 |
| Lymphocyte_percentage | OGTT0H | Weighted median | 12 | 0.298 | 0.048 | 4.45E-10 | Inverse variance weighted | 33.35787269 | 11 | 0.000 | -0.029 | 0.009 | 0.008 |
| Lymphocyte_percentage | OGTT0H | Inverse variance weighted | 12 | 0.258 | 0.061 | 2.48E-05 | Inverse variance weighted | 33.35787269 | 11 | 0.000 | -0.029 | 0.009 | 0.008 |
| Lymphocyte_percentage | OGTT0H | Simple mode | 12 | 0.313 | 0.067 | 7.00E-04 | Inverse variance weighted | 33.35787269 | 11 | 0.000 | -0.029 | 0.009 | 0.008 |
| Lymphocyte_percentage | OGTT0H | Weighted mode | 12 | 0.306 | 0.052 | 1.02E-04 | Inverse variance weighted | 33.35787269 | 11 | 0.000 | -0.029 | 0.009 | 0.008 |
| Neutrophil_percentage | OGTT0H | MR Egger | 13 | -0.374 | 0.049 | 9.60E-06 | Inverse variance weighted | 15.0566031 | 12 | 0.238 | 0.017 | 0.007 | 0.029 |
| Neutrophil_percentage | OGTT0H | Weighted median | 13 | -0.305 | 0.047 | 1.31E-10 | Inverse variance weighted | 15.0566031 | 12 | 0.238 | 0.017 | 0.007 | 0.029 |
| Neutrophil_percentage | OGTT0H | Inverse variance weighted | 13 | -0.285 | 0.037 | 2.19E-14 | Inverse variance weighted | 15.0566031 | 12 | 0.238 | 0.017 | 0.007 | 0.029 |
| Neutrophil_percentage | OGTT0H | Simple mode | 13 | -0.325 | 0.076 | 1.11E-03 | Inverse variance weighted | 15.0566031 | 12 | 0.238 | 0.017 | 0.007 | 0.029 |
| Neutrophil_percentage | OGTT0H | Weighted mode | 13 | -0.299 | 0.051 | 7.39E-05 | Inverse variance weighted | 15.0566031 | 12 | 0.238 | 0.017 | 0.007 | 0.029 |
| White_blood_cell | OGTT0H | MR Egger | 19 | -0.185 | 0.030 | 1.05E-05 | Inverse variance weighted | 25.24771879 | 18 | 0.118 | -0.007 | 0.005 | 0.167 |
| White_blood_cell | OGTT0H | Weighted median | 19 | -0.183 | 0.029 | 1.23E-10 | Inverse variance weighted | 25.24771879 | 18 | 0.118 | -0.007 | 0.005 | 0.167 |
| White_blood_cell | OGTT0H | Inverse variance weighted | 19 | -0.212 | 0.025 | 9.32E-18 | Inverse variance weighted | 25.24771879 | 18 | 0.118 | -0.007 | 0.005 | 0.167 |
| White_blood_cell | OGTT0H | Simple mode | 19 | -0.235 | 0.042 | 2.64E-05 | Inverse variance weighted | 25.24771879 | 18 | 0.118 | -0.007 | 0.005 | 0.167 |
| White_blood_cell | OGTT0H | Weighted mode | 19 | -0.184 | 0.029 | 6.33E-06 | Inverse variance weighted | 25.24771879 | 18 | 0.118 | -0.007 | 0.005 | 0.167 |
| Absolute_neutrophils | OGTT1H | MR Egger | 16 | 0.057 | 0.027 | 5.26E-02 | Inverse variance weighted | 15.63126803 | 15 | 0.407 | -0.009 | 0.005 | 0.079 |
| Absolute_neutrophils | OGTT1H | Weighted median | 16 | 0.041 | 0.028 | 1.46E-01 | Inverse variance weighted | 15.63126803 | 15 | 0.407 | -0.009 | 0.005 | 0.079 |
| Absolute_neutrophils | OGTT1H | Inverse variance weighted | 16 | 0.027 | 0.022 | 2.28E-01 | Inverse variance weighted | 15.63126803 | 15 | 0.407 | -0.009 | 0.005 | 0.079 |
| Absolute_neutrophils | OGTT1H | Simple mode | 16 | 0.030 | 0.037 | 4.32E-01 | Inverse variance weighted | 15.63126803 | 15 | 0.407 | -0.009 | 0.005 | 0.079 |
| Absolute_neutrophils | OGTT1H | Weighted mode | 16 | 0.044 | 0.030 | 1.66E-01 | Inverse variance weighted | 15.63126803 | 15 | 0.407 | -0.009 | 0.005 | 0.079 |
| Lymphocyte_percentage | OGTT1H | MR Egger | 12 | -0.039 | 0.066 | 5.66E-01 | MR Egger | 16.96886353 | 10 | 0.075 | -0.005 | 0.009 | 0.573 |
| Lymphocyte_percentage | OGTT1H | Weighted median | 12 | -0.065 | 0.048 | 1.77E-01 | MR Egger | 16.96886353 | 10 | 0.075 | -0.005 | 0.009 | 0.573 |
| Lymphocyte_percentage | OGTT1H | Inverse variance weighted | 12 | -0.068 | 0.044 | 1.24E-01 | MR Egger | 16.96886353 | 10 | 0.075 | -0.005 | 0.009 | 0.573 |
| Lymphocyte_percentage | OGTT1H | Simple mode | 12 | -0.035 | 0.069 | 6.20E-01 | MR Egger | 16.96886353 | 10 | 0.075 | -0.005 | 0.009 | 0.573 |
| Lymphocyte_percentage | OGTT1H | Weighted mode | 12 | -0.061 | 0.048 | 2.31E-01 | MR Egger | 16.96886353 | 10 | 0.075 | -0.005 | 0.009 | 0.573 |
| Neutrophil_percentage | OGTT1H | MR Egger | 13 | 0.052 | 0.055 | 3.65E-01 | MR Egger | 14.22135177 | 11 | 0.221 | -0.003 | 0.008 | 0.722 |

| | | | | | | | | | | | | | |
|---|---|---|---|---|---|---|---|---|---|---|---|---|---|
| Neutrophil_percentage | OGTT1H | Weighted median | 13 | 0.062 | 0.041 | 1.31E-01 | MR Egger | 14.22135177 | 11 | 0.221 | -0.003 | 0.008 | 0.722 |
| Neutrophil_percentage | OGTT1H | Inverse variance weighted | 13 | 0.037 | 0.036 | 3.02E-01 | MR Egger | 14.22135177 | 11 | 0.221 | -0.003 | 0.008 | 0.722 |
| Neutrophil_percentage | OGTT1H | Simple mode | 13 | 0.003 | 0.068 | 9.63E-01 | MR Egger | 14.22135177 | 11 | 0.221 | -0.003 | 0.008 | 0.722 |
| Neutrophil_percentage | OGTT1H | Weighted mode | 13 | 0.063 | 0.052 | 2.47E-01 | MR Egger | 14.22135177 | 11 | 0.221 | -0.003 | 0.008 | 0.722 |
| White_blood_cell | OGTT1H | MR Egger | 19 | 0.064 | 0.026 | 2.54E-02 | Inverse variance weighted | 20.17334312 | 18 | 0.323 | -0.009 | 0.004 | 0.046 |
| White_blood_cell | OGTT1H | Weighted median | 19 | 0.039 | 0.028 | 1.65E-01 | Inverse variance weighted | 20.17334312 | 18 | 0.323 | -0.009 | 0.004 | 0.046 |
| White_blood_cell | OGTT1H | Inverse variance weighted | 19 | 0.030 | 0.022 | 1.74E-01 | Inverse variance weighted | 20.17334312 | 18 | 0.323 | -0.009 | 0.004 | 0.046 |
| White_blood_cell | OGTT1H | Simple mode | 19 | 0.034 | 0.038 | 3.77E-01 | Inverse variance weighted | 20.17334312 | 18 | 0.323 | -0.009 | 0.004 | 0.046 |
| White_blood_cell | OGTT1H | Weighted mode | 19 | 0.045 | 0.026 | 1.07E-01 | Inverse variance weighted | 20.17334312 | 18 | 0.323 | -0.009 | 0.004 | 0.046 |
| Absolute_neutrophils | OGTT2H | MR Egger | 16 | -0.047 | 0.027 | 1.07E-01 | MR Egger | 12.9342055 | 14 | 0.532 | -0.004 | 0.005 | 0.457 |
| Absolute_neutrophils | OGTT2H | Weighted median | 16 | -0.031 | 0.030 | 3.04E-01 | MR Egger | 12.9342055 | 14 | 0.532 | -0.004 | 0.005 | 0.457 |
| Absolute_neutrophils | OGTT2H | Inverse variance weighted | 16 | -0.060 | 0.022 | 6.95E-03 | MR Egger | 12.9342055 | 14 | 0.532 | -0.004 | 0.005 | 0.457 |
| Absolute_neutrophils | OGTT2H | Simple mode | 16 | -0.062 | 0.041 | 1.49E-01 | MR Egger | 12.9342055 | 14 | 0.532 | -0.004 | 0.005 | 0.457 |
| Absolute_neutrophils | OGTT2H | Weighted mode | 16 | -0.033 | 0.029 | 2.72E-01 | MR Egger | 12.9342055 | 14 | 0.532 | -0.004 | 0.005 | 0.457 |
| Lymphocyte_percentage | OGTT2H | MR Egger | 12 | 0.098 | 0.057 | 1.19E-01 | Inverse variance weighted | 15.824248 | 11 | 0.148 | -0.013 | 0.008 | 0.129 |
| Lymphocyte_percentage | OGTT2H | Weighted median | 12 | 0.046 | 0.046 | 3.16E-01 | Inverse variance weighted | 15.824248 | 11 | 0.148 | -0.013 | 0.008 | 0.129 |
| Lymphocyte_percentage | OGTT2H | Inverse variance weighted | 12 | 0.029 | 0.042 | 4.98E-01 | Inverse variance weighted | 15.824248 | 11 | 0.148 | -0.013 | 0.008 | 0.129 |
| Lymphocyte_percentage | OGTT2H | Simple mode | 12 | 0.051 | 0.064 | 4.45E-01 | Inverse variance weighted | 15.824248 | 11 | 0.148 | -0.013 | 0.008 | 0.129 |
| Lymphocyte_percentage | OGTT2H | Weighted mode | 12 | 0.049 | 0.052 | 3.72E-01 | Inverse variance weighted | 15.824248 | 11 | 0.148 | -0.013 | 0.008 | 0.129 |
| Neutrophil_percentage | OGTT2H | MR Egger | 13 | -0.108 | 0.049 | 5.03E-02 | Inverse variance weighted | 12.41876009 | 12 | 0.413 | 0.010 | 0.007 | 0.159 |
| Neutrophil_percentage | OGTT2H | Weighted median | 13 | -0.044 | 0.045 | 3.29E-01 | Inverse variance weighted | 12.41876009 | 12 | 0.413 | 0.010 | 0.007 | 0.159 |
| Neutrophil_percentage | OGTT2H | Inverse variance weighted | 13 | -0.054 | 0.034 | 1.16E-01 | Inverse variance weighted | 12.41876009 | 12 | 0.413 | 0.010 | 0.007 | 0.159 |
| Neutrophil_percentage | OGTT2H | Simple mode | 13 | -0.045 | 0.063 | 4.89E-01 | Inverse variance weighted | 12.41876009 | 12 | 0.413 | 0.010 | 0.007 | 0.159 |
| Neutrophil_percentage | OGTT2H | Weighted mode | 13 | -0.041 | 0.046 | 3.83E-01 | Inverse variance weighted | 12.41876009 | 12 | 0.413 | 0.010 | 0.007 | 0.159 |
| White_blood_cell | OGTT2H | MR Egger | 19 | -0.045 | 0.026 | 1.02E-01 | MR Egger | 15.89452642 | 17 | 0.531 | -0.002 | 0.004 | 0.692 |
| White_blood_cell | OGTT2H | Weighted median | 19 | -0.033 | 0.029 | 2.47E-01 | MR Egger | 15.89452642 | 17 | 0.531 | -0.002 | 0.004 | 0.692 |
| White_blood_cell | OGTT2H | Inverse variance weighted | 19 | -0.052 | 0.021 | 1.30E-02 | MR Egger | 15.89452642 | 17 | 0.531 | -0.002 | 0.004 | 0.692 |
| White_blood_cell | OGTT2H | Simple mode | 19 | -0.076 | 0.041 | 7.92E-02 | MR Egger | 15.89452642 | 17 | 0.531 | -0.002 | 0.004 | 0.692 |
| White_blood_cell | OGTT2H | Weighted mode | 19 | -0.042 | 0.027 | 1.36E-01 | MR Egger | 15.89452642 | 17 | 0.531 | -0.002 | 0.004 | 0.692 |

Method_het and Q_pval refer to the model of heterogeneity test and p value of statistic test; egger_intercept, pleio_se,pleio_p refer to the intercept,se and P value of pleiotropy test.

### Supplementary Figures

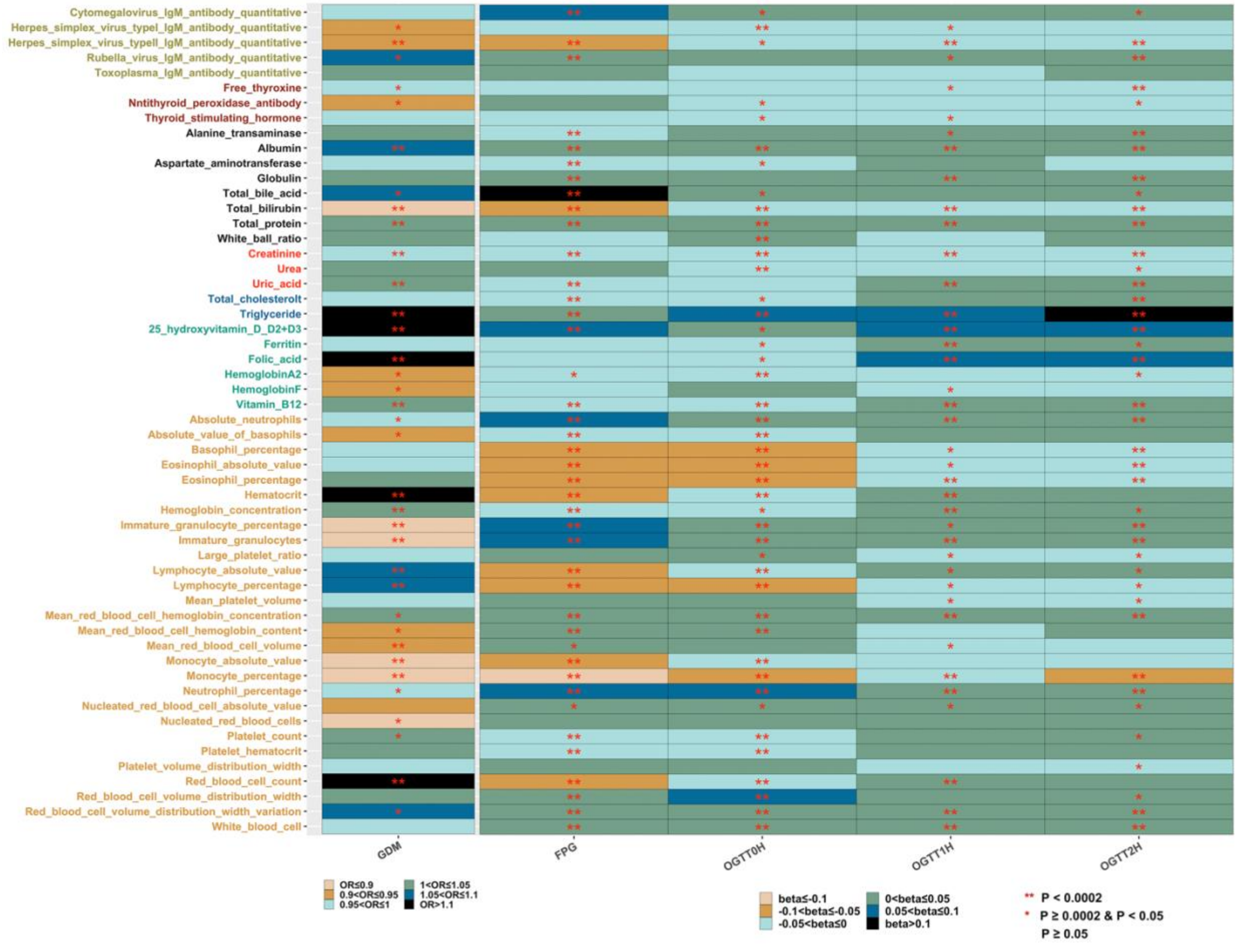

**ESM Fig. 1 Multiple regression analyses of GDM and the four glycemic traits.**
Standardized regression coefficients are displayed. The color of the vertical coordinate represents the categories of the biomarkers according to ESM Table 2, grass green: Infection; red-brown: Thyroid function; black: Liver function; red: Kidney function; blue: Blood lipid; cyan: Blood anemia; yellow: Blood routine. Color in square for qualitative variables indicates OR, for quantitative variables indicates effect size. The number of * sign in the square represents statistical test significance, bonferroni-adjusted threashold $\alpha$ is 0.0001(0.05/55/5).

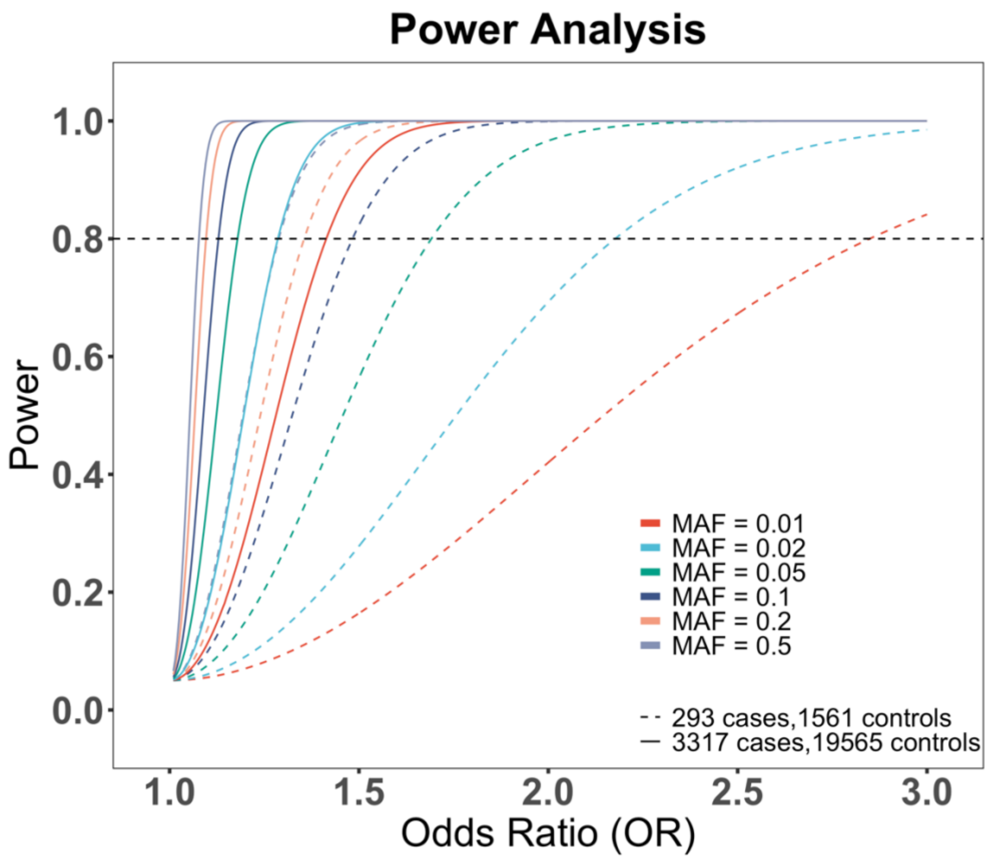

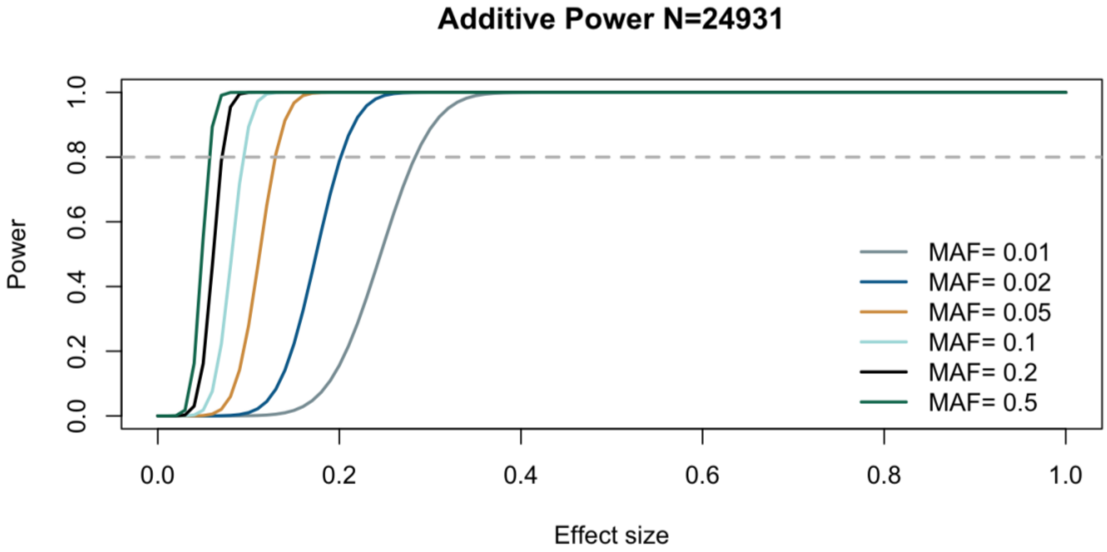

**ESM Fig. 2 Power analysis of the genome-wide association analysis.**

Top: Power analysis for GWAS of GDM susceptibility (3,317 cases versus 19,565 controls) mean this study and GWAS of GDM susceptibility in BIGCS replication cohort (293 cases versus 1,561 controls). Significance threshold P value < $5\times10^{-8}$. Bottom: Power analysis for GWAS of the four glycemic traits. Significance threshold P value < $5\times10^{-8}$. MAF: minor allele frequency.

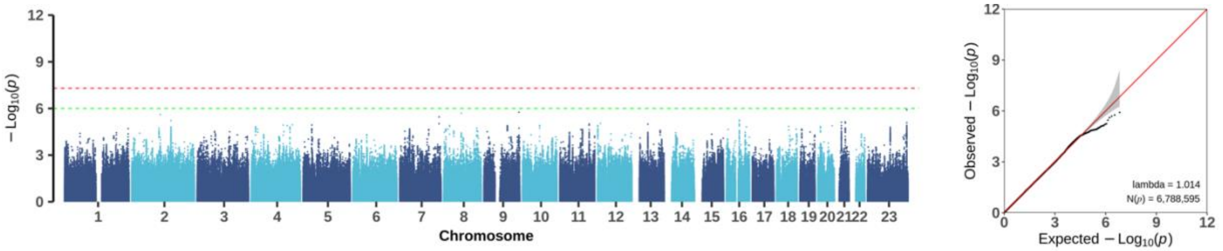

**ESM Fig. 3 Manhattan plot of GDM case with medication or not (case/control=419/2898).**

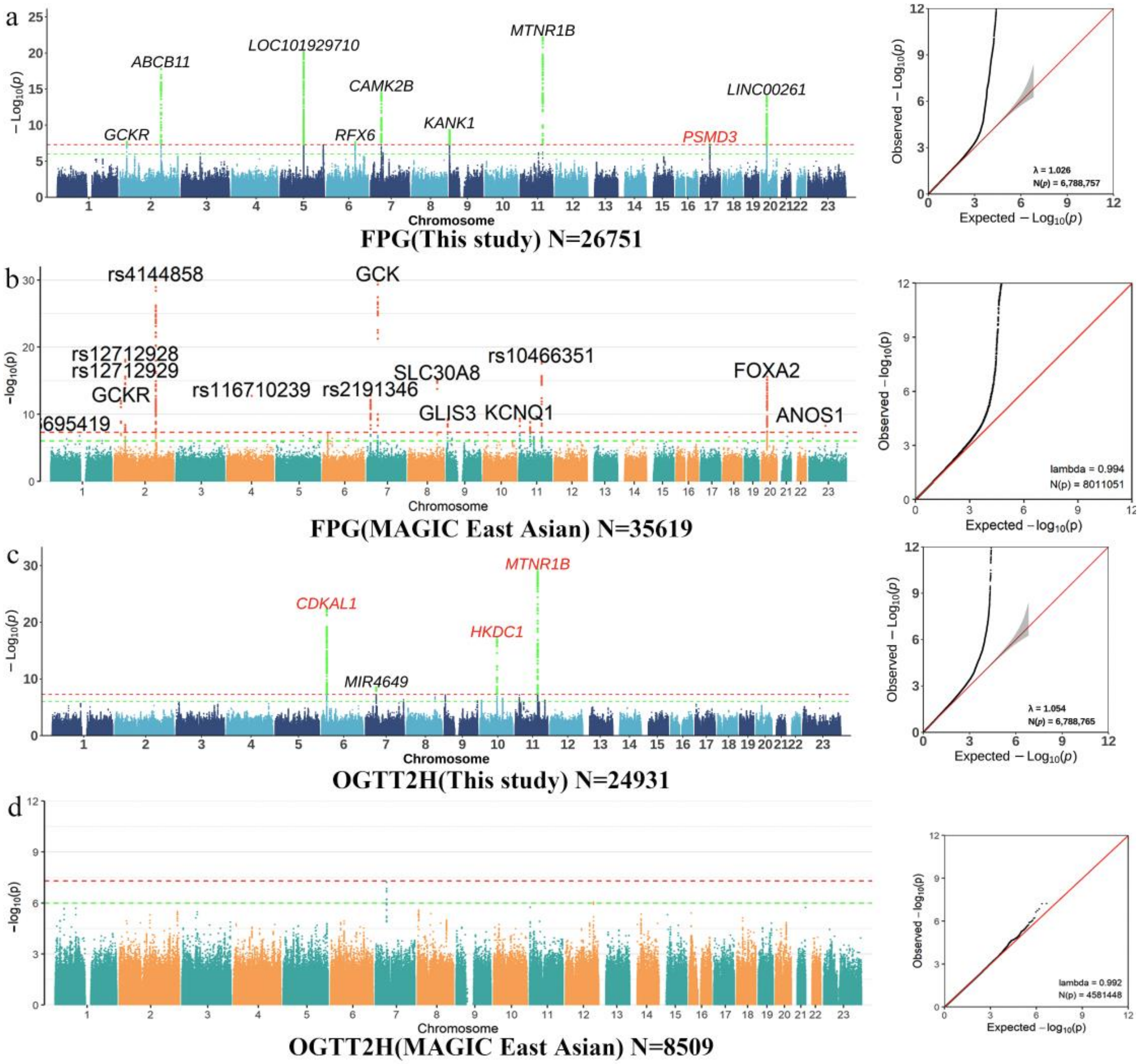

**ESM Fig. 4 Comparison of the genome-wide association analysis for FPG and OGTT2H between MAGIC East Asian and our study.**

For panel a and c, results from our study, newly identified loci are denoted by red signals, while loci with established knowledge are represented in black. For panel B and D, the loci were know and came from MAGIC East Asian OGTT2H and FPG GWAS summary, which were downloaded from https://magicinvestigators.org/downloads/.

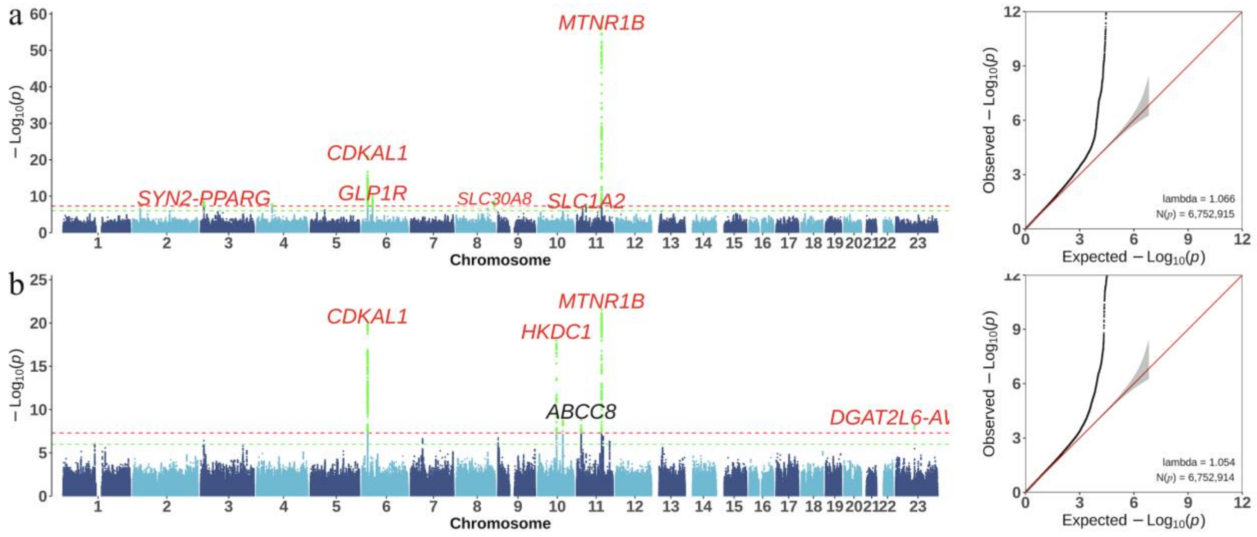

**ESM Fig. 5 Manhattan and qq plot of OGTT1H(a) and OGTT2H(b) with age,BMI, gestational week of OGTT and FPG as covariants.**

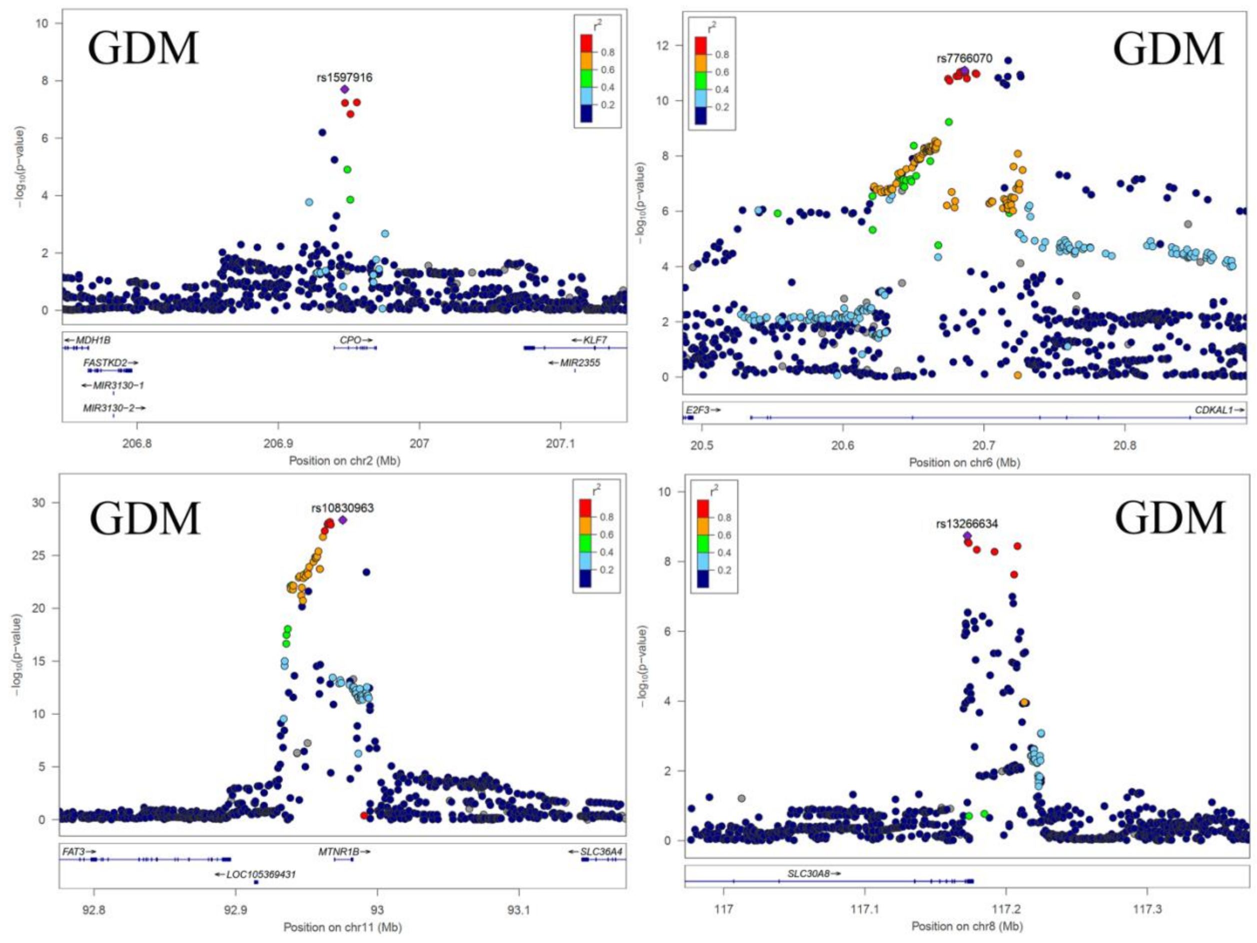

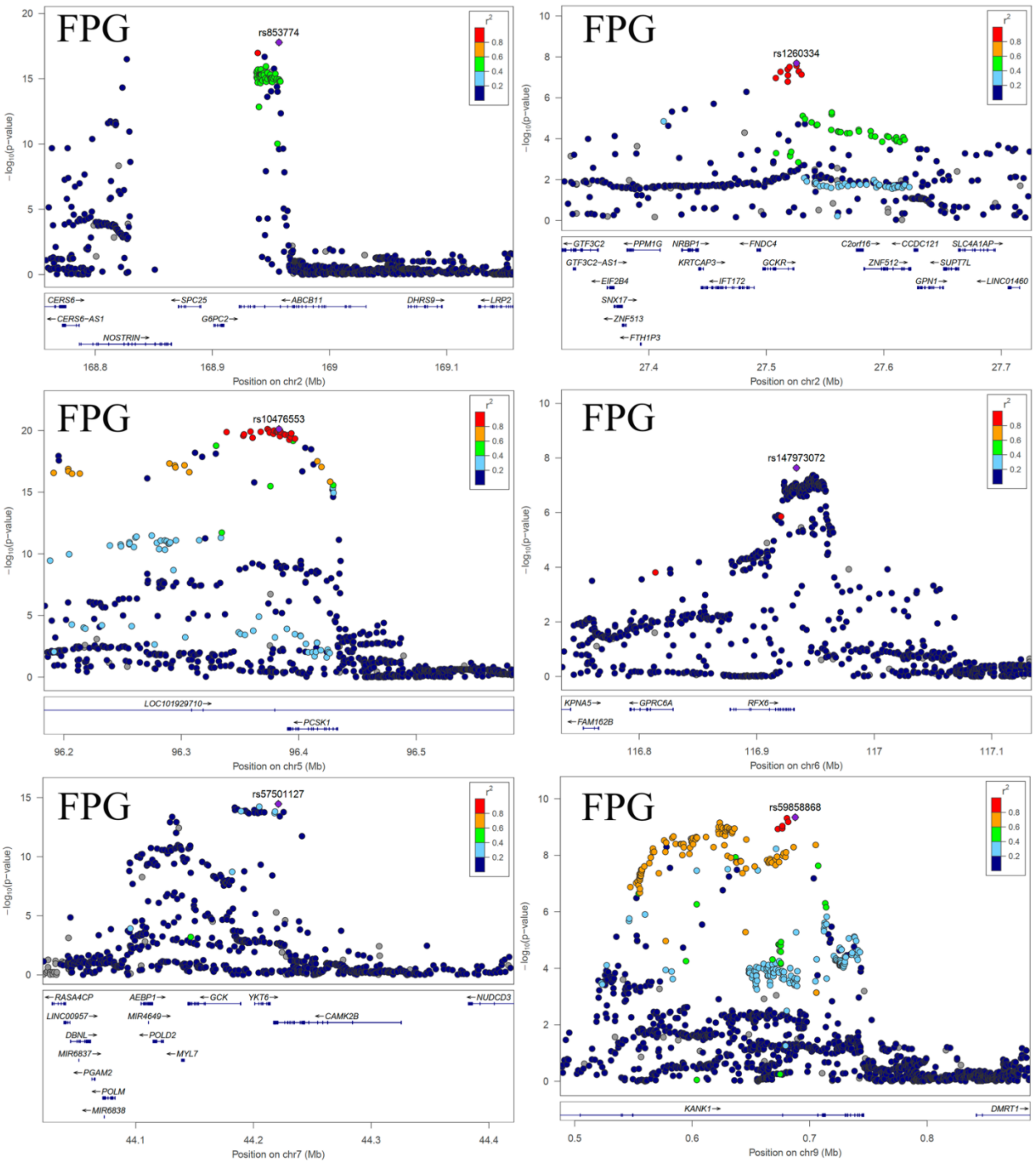
FPG
rs853774
−log10(p−value)
r2
0.8
0.6
0.4
0.2
CERS6→
←CERS6−AS1
NOSTRIN→
←SPC25
G6PC2→
←ABCB11
DHRS9→
←LRP2
168.8
168.9
169
169.1
Position on chr2 (Mb)
FPG
rs1260334
←GTF3C2
GTF3C2−AS1→
←EIF2B4
SNX17→
←ZNF513
←FTH1P3
←PPM1G
NRBP1→
KRTCAP3→
←IFT172
←FNDC4
GCKR→
C2orf16→
ZNF512→
←CCDC121
GPN1→
SLC4A1AP→
←SUPT7L
←LINC01460
27.4
27.5
27.6
27.7
Position on chr2 (Mb)
FPG
rs10476553
LOC101929710→
←PCSK1
96.2
96.3
96.4
96.5
Position on chr5 (Mb)
FPG
rs147973072
KPNA5→
←FAM162B
←GPRC6A
RFX6→
116.8
116.9
117
117.1
Position on chr6 (Mb)
FPG
rs57501127
←RASA4CP
LINC00957→
DBNL→
MIR6837→
←PGAM2
←POLM
←MIR6838
AEBP1→
MIR4649→
←POLD2
←MYL7
←GCK
YKT6→
←CAMK2B
←NUDCD3
44.1
44.2
44.3
44.4
Position on chr7 (Mb)
FPG
rs59858868
KANK1→
DMRT1→
0.5
0.6
0.7
0.8
Position on chr9 (Mb)

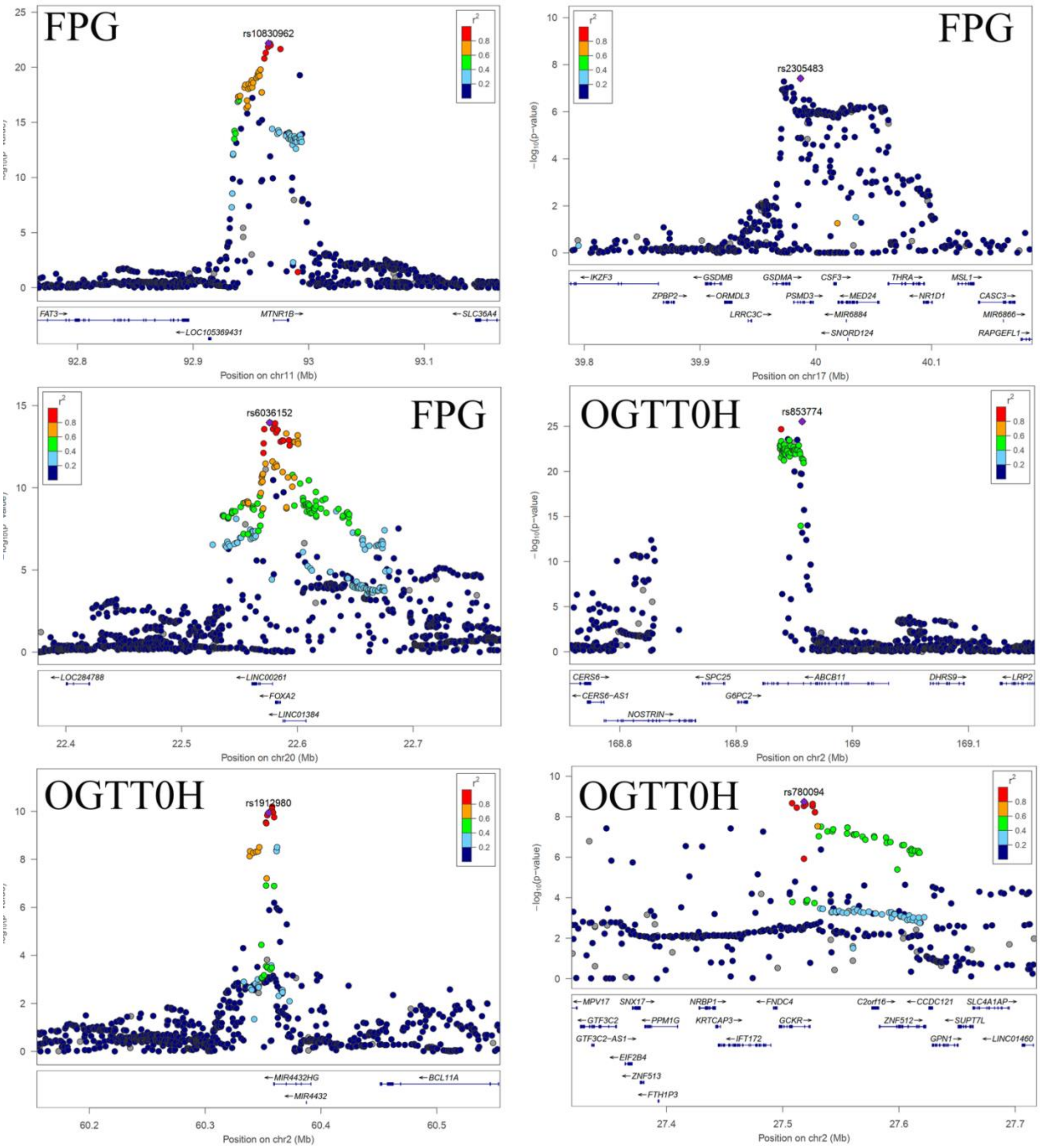
FPG
rs10830962
FAT3→
MTNR1B→
←SLC36A4
←LOC105369431
Position on chr11 (Mb)
FPG
rs2305483
−log10(p−value)
←IKZF3
←GSDMB
GSDMA→
CSF3→
THRA→
MSL1→
ZPBP2→
←ORMDL3
PSMD3→
←MED24
←NR1D1
CASC3→
LRRC3C→
←MIR6884
MIR6866→
←SNORD124
RAPGEFL1→
Position on chr17 (Mb)
FPG
rs6036152
←LOC284788
←LINC00261
←FOXA2
←LINC01384
Position on chr20 (Mb)
OGTT0H
rs853774
−log10(p−value)
CERS6→
←SPC25
←ABCB11
DHRS9→
←LRP2
←CERS6−AS1
G6PC2→
NOSTRIN→
Position on chr2 (Mb)
OGTT0H
rs1912980
←MIR4432HG
←BCL11A
←MIR4432
Position on chr2 (Mb)
OGTT0H
rs780094
−log10(p−value)
←MPV17
SNX17→
NRBP1→
←FNDC4
C2orf16→
←CCDC121
SLC4A1AP→
←GTF3C2
←PPM1G
KRTCAP3→
GCKR→
ZNF512→
←SUPT7L
GTF3C2−AS1→
←IFT172
GPN1→
←LINC01460
←EIF2B4
←ZNF513
←FTH1P3
Position on chr2 (Mb)

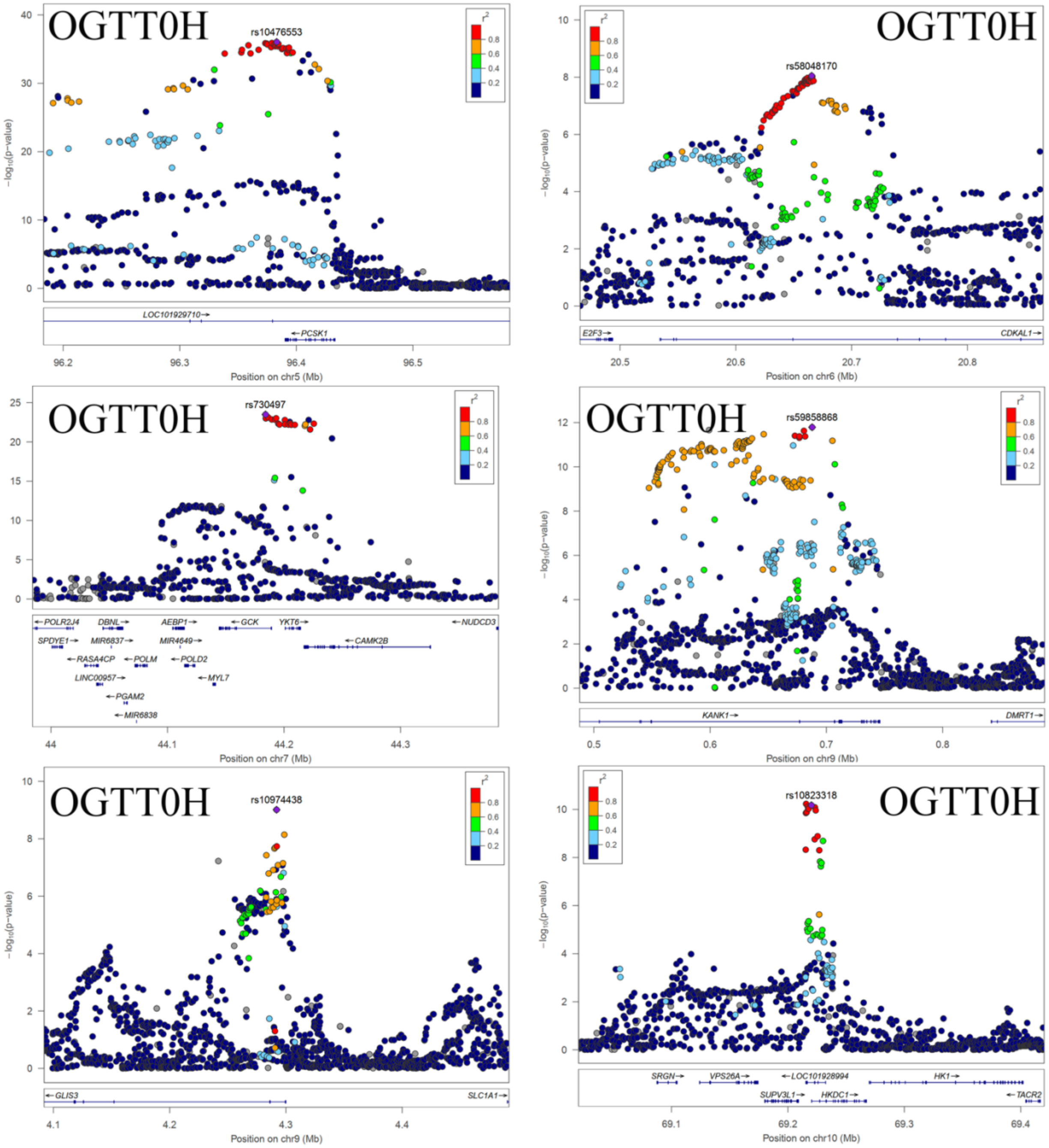

OGTT0H
rs10476553
LOC101929710→
←PCSK1
Position on chr5 (Mb)
OGTT0H
rs58048170
E2F3→
CDKAL1→
Position on chr6 (Mb)
OGTT0H
rs730497
←POLR2J4
DBNL→
AEBP1→
←GCK
YKT6→
←NUDCD3
SPDYE1→
MIR6837→
MIR4649→
←CAMK2B
←RASA4CP
←POLM
←POLD2
LINC00957→
←MYL7
←PGAM2
←MIR6838
Position on chr7 (Mb)
OGTT0H
rs59858868
KANK1→
DMRT1→
Position on chr9 (Mb)
OGTT0H
rs10974438
←GLIS3
SLC1A1→
Position on chr9 (Mb)
OGTT0H
rs10823318
SRGN→
VPS26A→
←LOC101928994
HK1→
SUPV3L1→
HKDC1→
←TACR2
Position on chr10 (Mb)
−log10(p−value)
r2

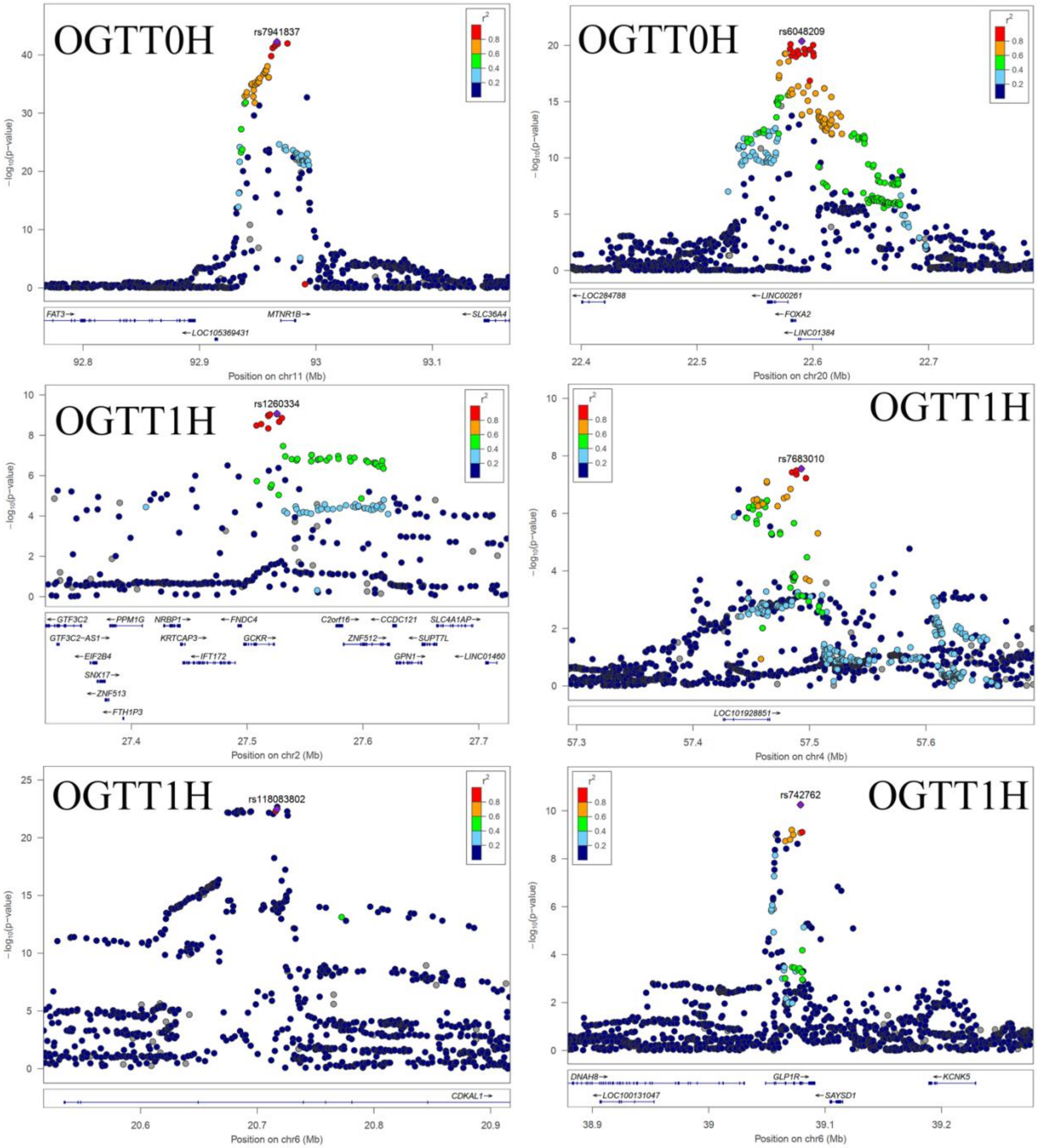
OGTT0H
rs7941837
−log10(p−value)
FAT3→
MTNR1B→
←SLC36A4
←LOC105369431
Position on chr11 (Mb)
OGTT0H
rs6048209
←LOC284788
←LINC00261
←FOXA2
←LINC01384
Position on chr20 (Mb)
OGTT1H
rs1260334
←GTF3C2
←PPM1G
NRBP1→
←FNDC4
C2orf16→
←CCDC121
SLC4A1AP→
GTF3C2−AS1→
KRTCAP3→
GCKR→
ZNF512→
←SUPT7L
←EIF2B4
←IFT172
GPN1→
←LINC01460
SNX17→
←ZNF513
←FTH1P3
Position on chr2 (Mb)
OGTT1H
rs7683010
LOC101928851→
Position on chr4 (Mb)
OGTT1H
rs118083802
CDKAL1→
Position on chr6 (Mb)
OGTT1H
rs742762
DNAH8→
GLP1R→
←KCNK5
←LOC100131047
←SAYSD1
Position on chr6 (Mb)

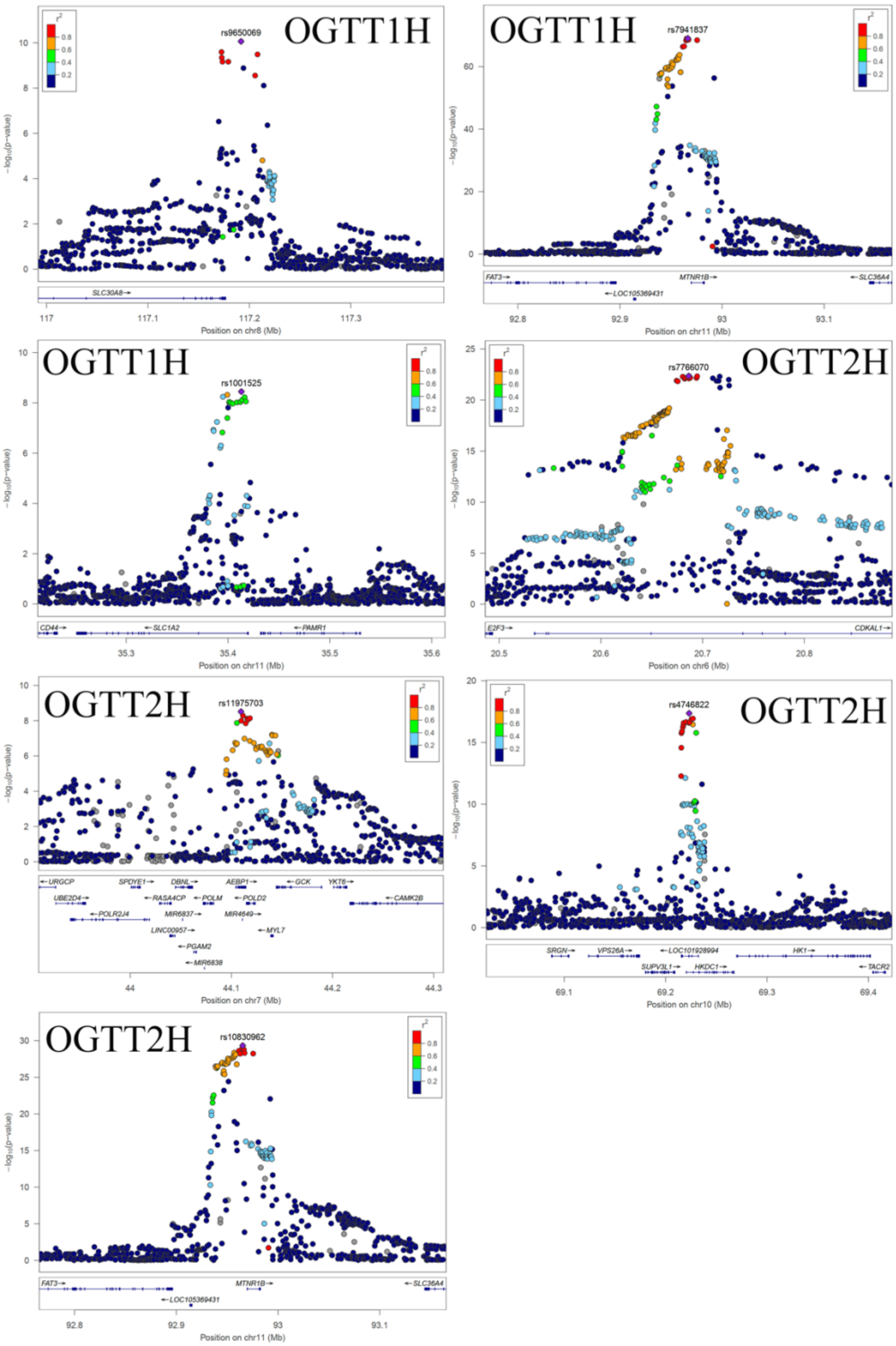

**ESM Fig. 6 Locuszoom plot of genome-wide significant loci associated with the seven traits investigated in the study.**

For all the 35 lead SNPs in ESM Table 8, a regional plot showing the P-value and the LD $r^2$ of SNPs located in the upstream and downstream 500kbp flanking region are demonstrated using the Locuszoom software.

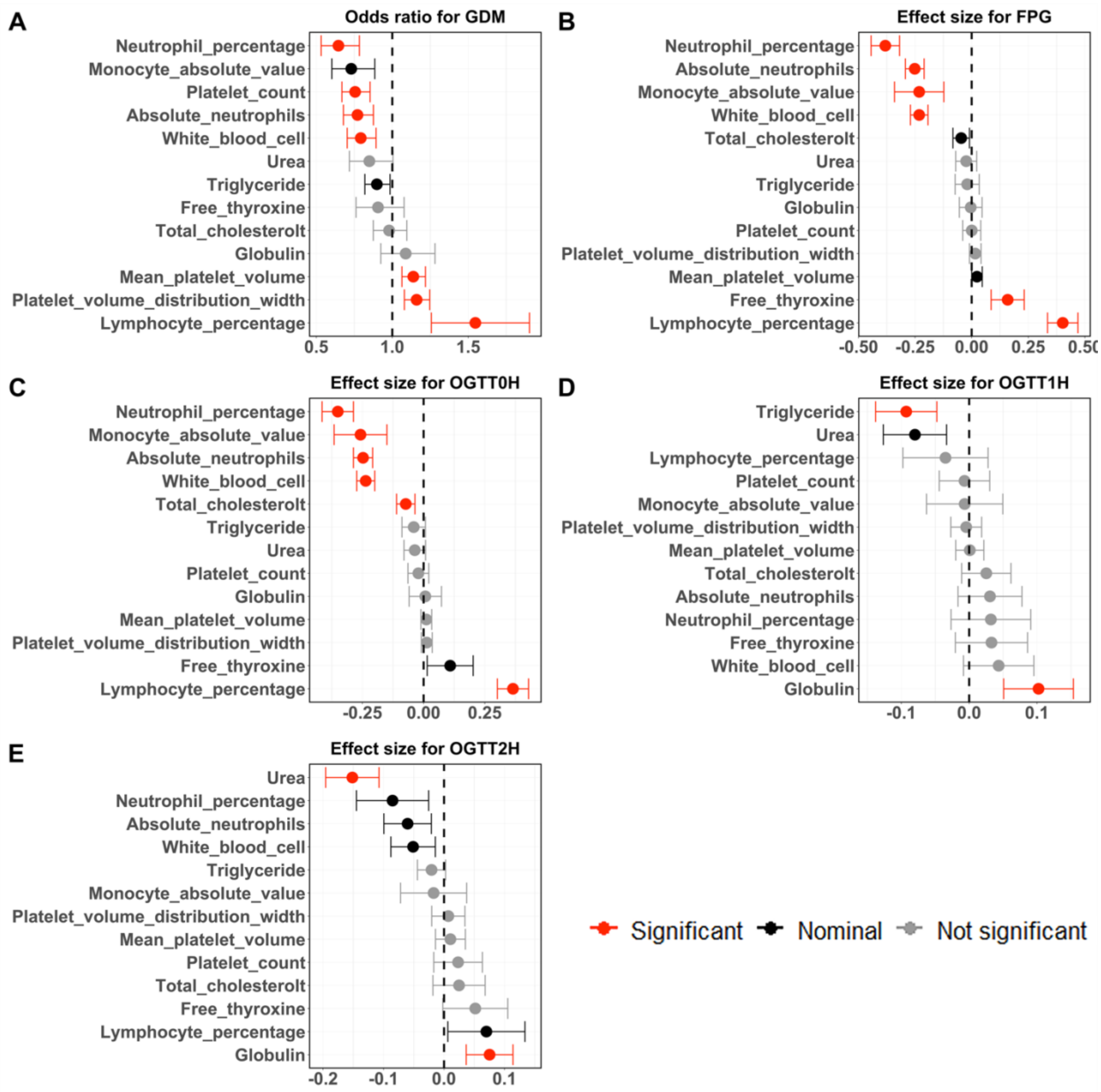

**ESM Fig. 7 Effects of biomarkers on GDM and four glycemic traits by mendelian randomization.**

All the 55 biomarkers collected in the pregnancy screening were analysed towards the GDM and the four quantitative glycemic traits.

Significant causal associations were marked in red where the following two criteria were met (1) P-values of inverse variance weight (IVW) was less than the bonferroni correction threshold ($\alpha = 0.05/51/5$) and (2) either the association showed no evidence of heterogeneity ($p > 0.05$) and horizontal pleiotropy ($p > 0.05$), or one of the MR Egger, Simple mode, Weighted median, Weighted mode corrected estimates also have a $p < 0.05$). Nominal significant causal association were marked in black using the similar criteria except that in criterion, P-value of IVW was less than 0.05 rather than less than bonferroni correction threshold. Associations that did not meet the significant or the nominal significant criteria were marked in gray.

Only biomarkers that demosntrate statistical significance after Bonferroni correction in at least one of the exposure-outcome analyses were included in the plot while results for all the biomarkers were presented in ESM Fig. 9.

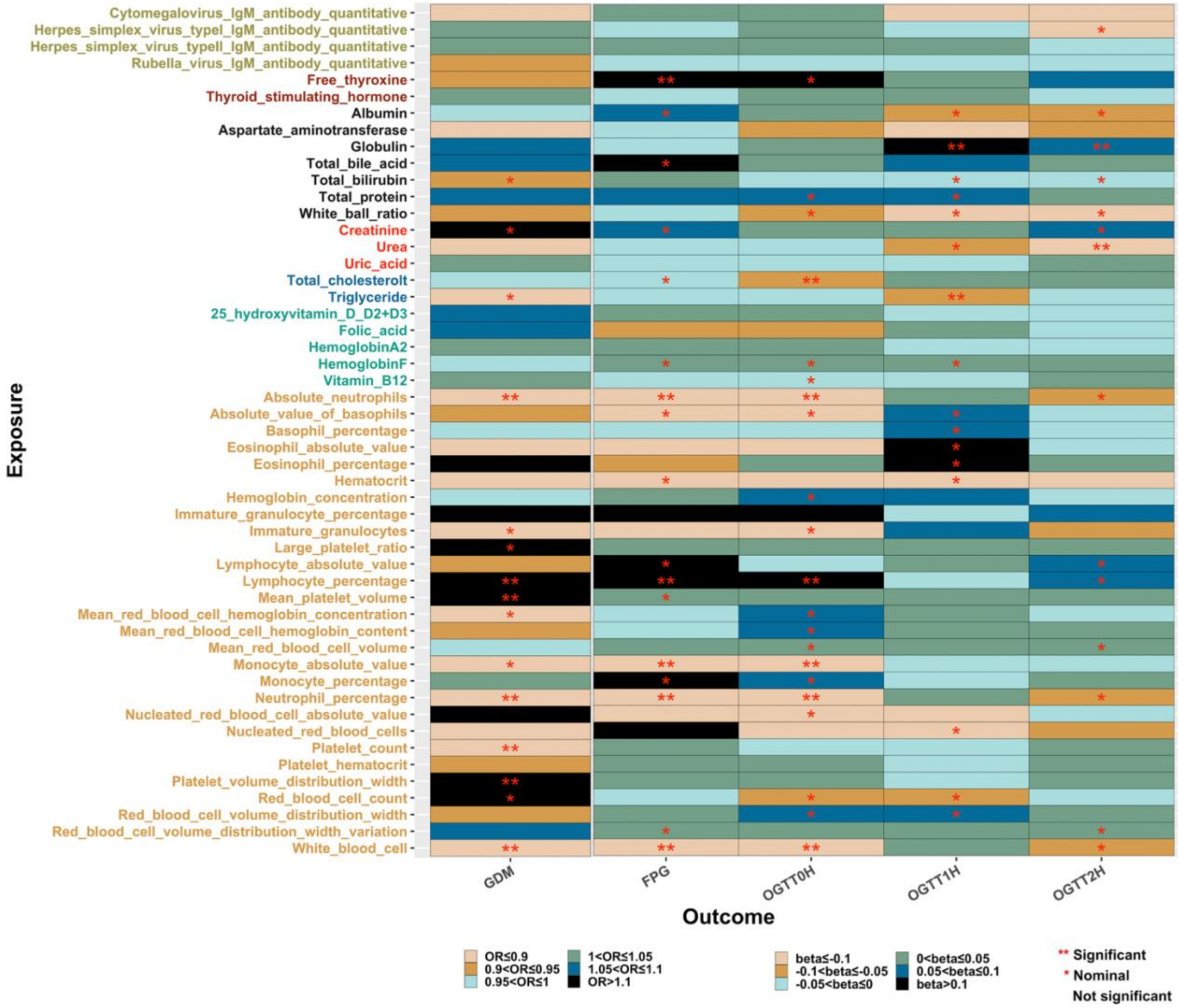

**ESM Fig. 8 Effects of all 51 biomarkers on GDM and glycemic traits by mendelian randomization.**

The rule for representing the color of the vertical coordinate, the color in the square was the same as for ESM Fig. 2. Square color indicate effect size of IVW, and the number of * sign in the square represents statistical significance, the rule of significance was the same as ESM Fig. 6, bonfferoni correction $\alpha=0.05/51/5$.

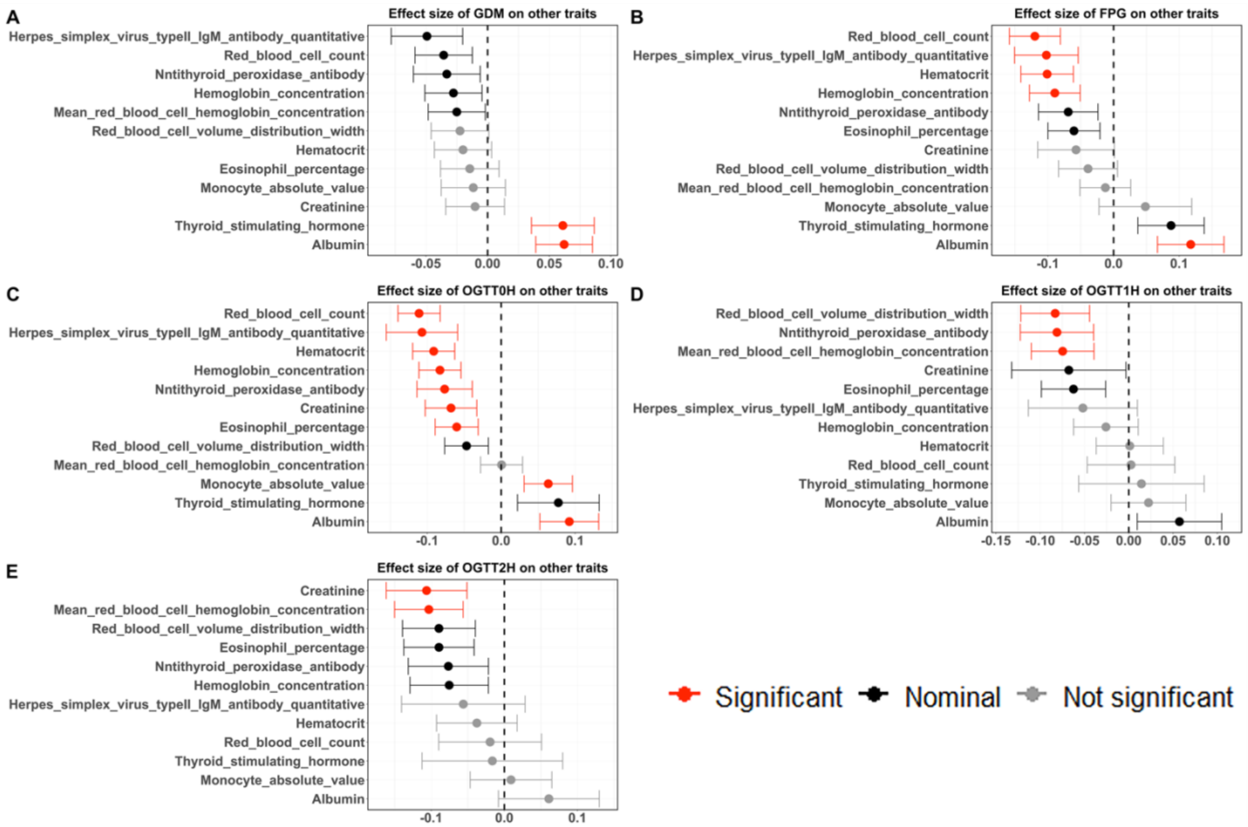

**ESM Fig. 9 Effects of GDM and glycemic traits on the biomarkers by Mendelian randomization.**

The analysis was conducted between all the 55 biomarkers collected in the pregnancy screening and the GDM occurrence as well as the 4 quantitative glycemic traits.

The definition of significance was the same as ESM Fig. 7, bonferroni correction $\alpha$=0.05/55/5.

Only biomarkers that demosntrate statistical significance after Bonferroni correction in at least one of the exposure-outcome analyses were included in the plot while results for all the biomarkers were presented in ESM Fig. 11.

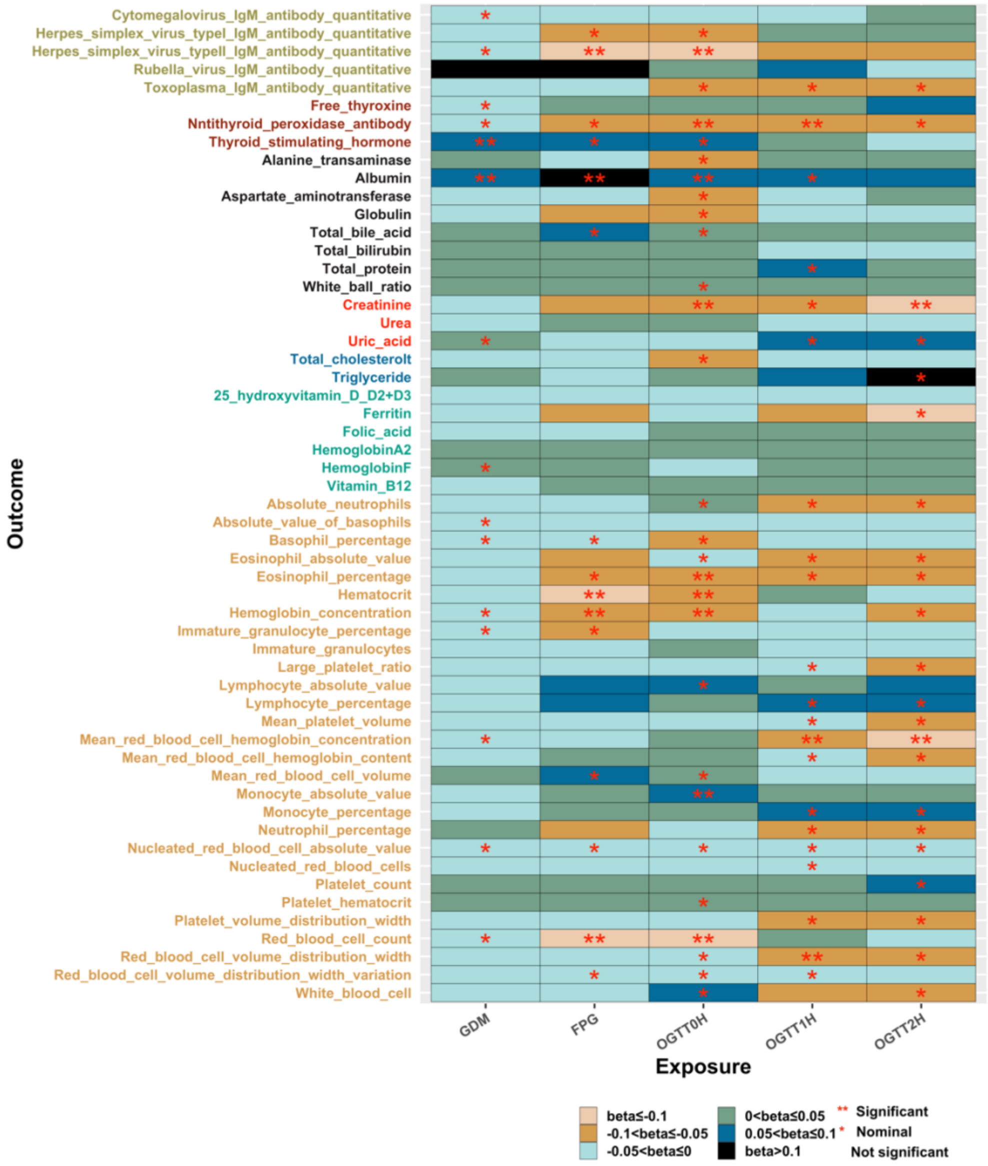

**ESM Fig. 10 Effects of GDM and glycemic traits on all the** 55 **biomarkers by Mendelian randomization.**

The rule for representing the color of the vertical coordinate, the color in the square was the same as for ESM Fig. 2. Square color indicate effect size of IVW, and the number of * sign in the square represents statistical significance, the definition of significance was the same as ESM Fig. 6, bonferroni correction $\alpha$=0.05/55/5.

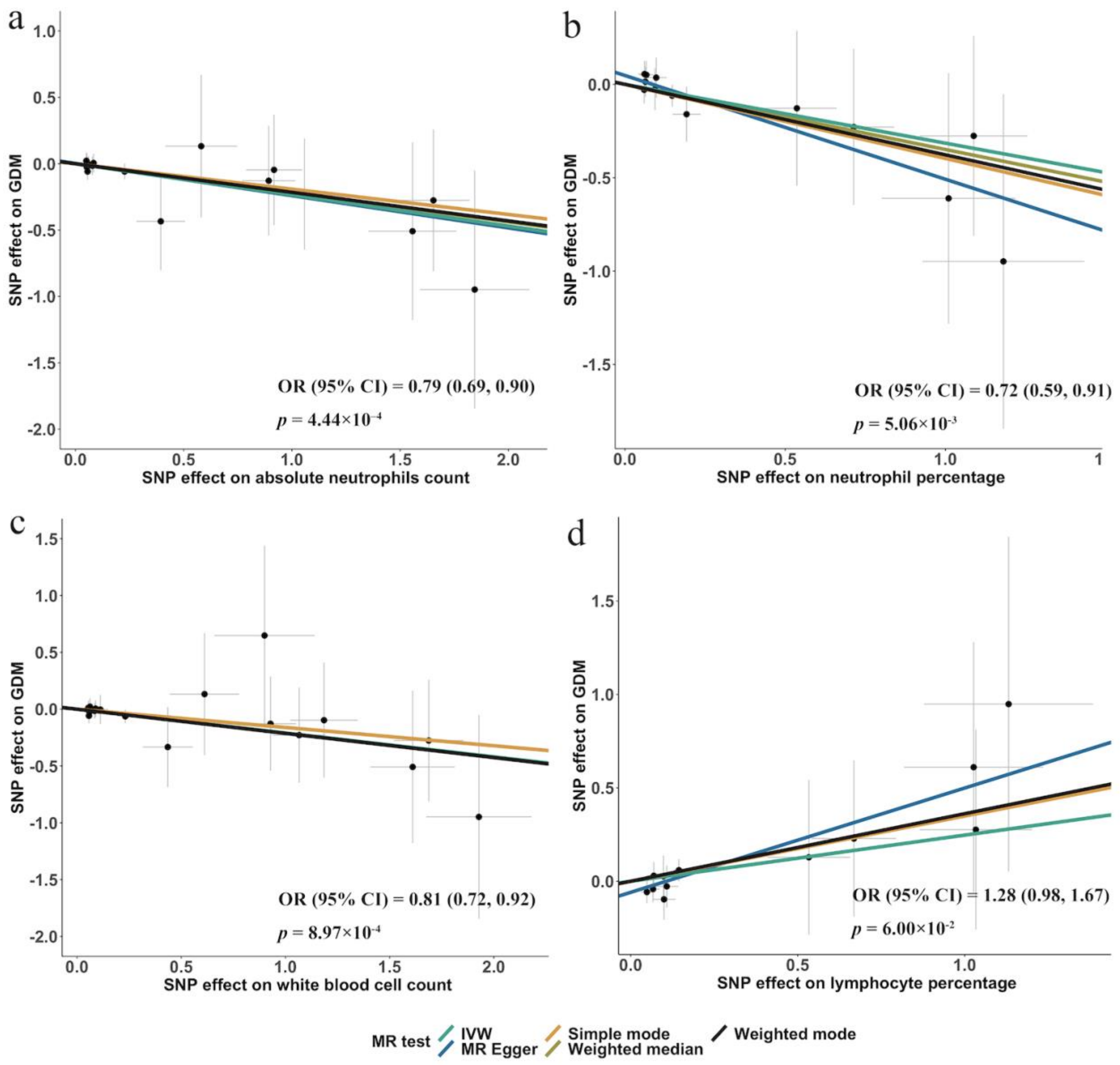

**ESM Fig. 11 Scatter plot of mendelian randomization analysis results with 4 biomarkers as the exposure and their effect on GDM with PLUS cohort meta data.**

Panel a-d presents the results of the mendelian randomization analyses using absolute neutrophils, neutrophil percentage, white blood cell and lymphocyte percentage as exposure respectively. The effect size and P value of inverse variance weighted showed in figure and the slope of the regression line in each panel indicates the direction of the effect of the exposure on GDM, with a positive slope representing a positive effect and a negative slope indicating a negative effect.
